# Local activity alterations in autism spectrum disorder correlate with neurotransmitter properties and ketamine induced brain changes

**DOI:** 10.1101/2024.10.20.24315801

**Authors:** Pascal Grumbach, Jan Kasper, Joerg F. Hipp, Anna Forsyth, Sofie L. Valk, Suresh Muthukumaraswamy, Simon B. Eickhoff, Leonhard Schilbach, Juergen Dukart

## Abstract

Autism spectrum disorder (ASD) is a neurodevelopmental condition associated with altered resting-state brain function. An increased excitation-inhibition (E/I) ratio is discussed as a potential pathomechanism but in-vivo evidence of disturbed neurotransmission underlying these functional alterations remains scarce. We compared rs-fMRI local activity (LCOR) between ASD (N=405, N=395) and neurotypical controls (N=473, N=474) in two independent cohorts (ABIDE1 and ABIDE2). We then tested how these LCOR alterations co-localize with specific neurotransmitter systems derived from nuclear imaging and compared them with E/I changes induced by GABAergic (midazolam) and glutamatergic medication (ketamine). Across both cohorts, ASD subjects consistently exhibited reduced LCOR, particularly in higher-order default mode network nodes, alongside increases in bilateral temporal regions, the cerebellum, and brainstem. These LCOR alterations negatively co-localized with dopaminergic (D1, D2, DAT), glutamatergic (NMDA, mGluR5), GABAergic (GABAa) and cholinergic neurotransmission (VAChT). The NMDA-antagonist ketamine, but not GABAa-potentiator midazolam, induced LCOR changes which co-localize with D1, NMDA and GABAa receptors, thereby resembling alterations observed in ASD. We find consistent local activity alterations in ASD to be spatially associated with several major neurotransmitter systems. NMDA-antagonist ketamine induced neurochemical changes similar to ASD-related alterations, supporting the notion that pharmacological modulation of the E/I balance in healthy individuals can induce ASD-like functional brain changes. These findings provide novel insights into neurophysiological mechanisms underlying ASD.

**One Sentence Summary:** Local activity alterations in ASD co-localize with glutamatergic and GABAergic neurotransmission and were similar to ketamine-induced brain changes.

## INTRODUCTION

Autism spectrum disorder (ASD) is a neurodevelopmental condition primarily characterized by deficits in communication, impaired social interaction and presence of stereotyped and repetitive behavior. Diagnosis is based on behavior and frequently complicated by the heterogeneity of symptoms as well as the overlap with other psychiatric conditions^1,2^. The underlying neurobiological mechanisms of ASD remain poorly understood, reliable biomarkers are lacking, and as a result, an effective pharmacological treatment model has yet to be established^3^.

Resting-state functional magnetic resonance imaging (rs-fMRI) has the potential to offer easily applicable, objective neural biomarkers to assist traditional clinical diagnosis of ASD. Therefore, numerous neuroimaging studies have investigated resting-state functional alterations in individuals with ASD compared to typically developed controls (TD). Despite variability in these findings, potentially due to the use of different measures, meta-analyses have identified consistent patterns of both functional hyperactivity and -connectivity in the cerebellum, temporal and sensorimotor areas, alongside with hypoactivity and -connectivity within default mode network (DMN) nodes - a network important for self-referential processing and social cognition^4–11^. To date, little is known about specific neurotransmitter properties underlying this functional reorganization in ASD and how these alterations relate to specific symptom domains^12–14^. Emerging evidence has implicated the dopaminergic modulation of motor pathways in manifestation of stereotyped behavior^15,16^. Furthermore, increased serotonin reuptake has been linked to impaired social behavior and sensory development in a subset of children with ASD^17^.

Most prominent, alterations in glutamatergic neurotransmission have been proposed as pivotal in the pathophysiology of ASD^18,19^. For example, Galineau et al. (2022) reported an overall increased density of the metabotropic glutamate receptor mGluR5 in male adults with ASD^20^. Several studies support the hypothesis that N-Methyl-D-Aspartate (NMDA) receptor dysfunction contributes to ASD symptoms indicating that these symptoms can be improved by the NMDA receptor agonist D-Cycloserine^18,21^. Moreover, Siegel-Ramsey et al. (2021) found a negative association between increased glutamate concentrations in the dorsal anterior cingulate cortex (ACC) and reduced functional connectivity between the dorsal ACC and insular, limbic and parietal regions in male participants with ASD^22^. Findings of altered glutamate neurotransmission, coupled with evidence of GABAergic (γ-aminobutyric acid) deficit and synaptic dysregulation in ASD^23,24^, have converged to support the hypothesis of an elevated excitation/inhibition (E/I) ratio as a potential molecular model for ASD^25,26^. Despite an increasing body of evidence identifying ASD-related molecular and functional alterations, the relationship between both types of changes remains poorly understood. It also remains uncertain whether pharmacologically induced E/I modulation as elicited by glutamatergic medication ketamine and GABAergic medication midazolam in healthy subjects results in macroscale functional reorganization akin to that observed in ASD^27–29^.

The aim of the present study was to better understand the neurochemical basis of functional alterations in ASD by probing for their co-localization with *in-vivo* derived neurotransmitter information. We further test for similarity of ASD-related alterations with ketamine and midazolam induced functional brain changes. We hypothesize that (1) individuals with ASD show consistent local hyper- and hypoactivity in sensorimotor and DMN areas that co-localize with the spatial distribution of specific neurotransmitter and that (2) ketamine and midazolam induced neurochemical patterns are similar to these ASD-related alterations. More specifically, based on evidence from previous studies, we anticipate the strongest co-localizations with glutamatergic, GABAergic, serotonergic and dopaminergic neurotransmission^13,30^. To improve the robustness of our findings, all hypotheses were initially tested using the ABIDE1 dataset and subsequently validated in the independent ABIDE2 dataset^31^.

## RESULTS

### ASD subjects across datasets differ regarding age and symptom severity

In both datasets, individuals with ASD and TD controls did not differ in terms of age (Table 1). However, TD controls included a significantly higher proportion of female subjects. In ABIDE2, TD controls showed higher FIQ values as compared to ASD, whereas there was no FIQ difference in ABIDE1. Moreover, individuals with ASD showed more head motion during image acquisition than TD controls. ASD participants in ABIDE1 were older and exhibited a higher severity of autistic symptoms as compared to those in ABIDE2.

**Table 1.**
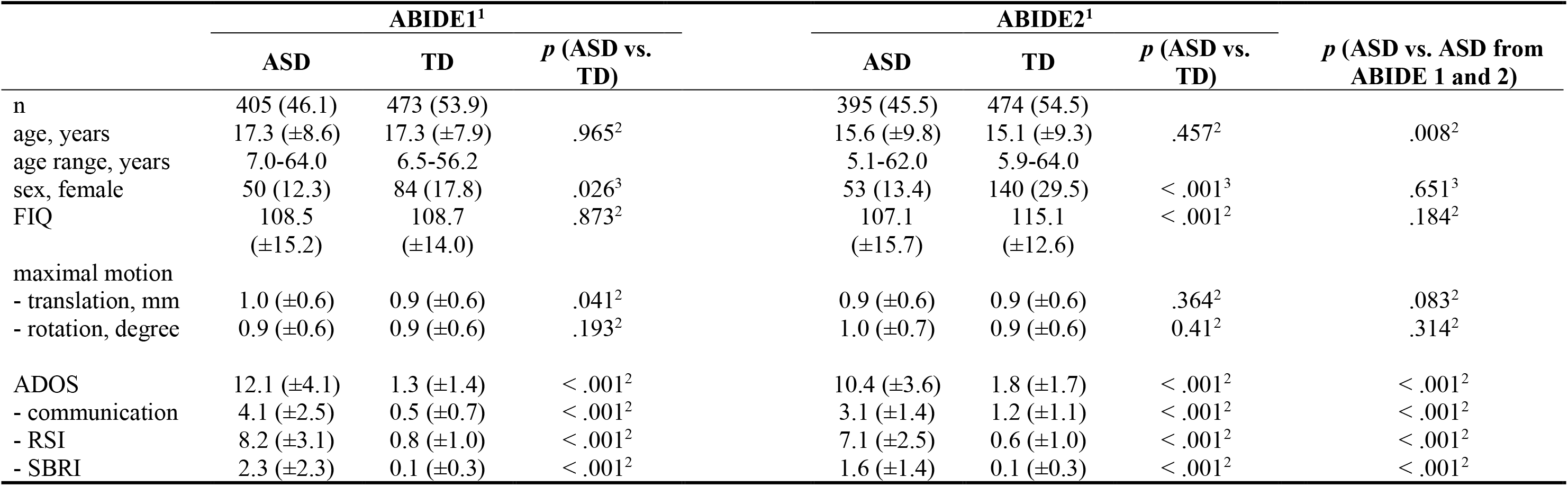
Demographic and clinical characteristics of the ABIDE1 and ABIDE2 cohorts. ^1^Numbers present either absolute numbers plus percent or mean plus standard deviation, ^2^*t*-test (two-tailed), ^3^*χ^2^*-test (two-tailed), ASD = autism spectrum disorder; TD = typically developed controls; ABIDE = Autism Brain Imaging Data Exchange; FIQ = full scale IQ; ADOS = Autism Diagnostic Observation Schedule; RSI = reciprocal social interaction; SBRI = stereotyped behaviors and restricted interests; maximal available clinical data for ABIDE-I ADOS (n = 300 ASD, 28 TD), for ABIDE-II ADOS (n = 238 ASD, 42 TD).

### Consistent local activity alterations in ASD

First, we tested for ASD-related alterations in local synchronization (LCOR) compared to TD controls. In whole-brain voxel-wise analyses in the ABIDE1 dataset, we found a pattern of LCOR reductions in ASD in brain regions comprising DMN hubs (bilateral posterior cingulate cortex (PCC), precuneus, frontal pole, frontal medial cortex), ACC, paracingulate gyrus and precentral gyrus as well as right hemispheric temporal pole, insular and opercular cortex (Figure 1A, Supplementary Table S1-S2). Increases in LCOR were observed in ASD in bilateral temporal regions, the cerebellum and right lateral occipital cortex and angular gyrus. The decrease but not the increase findings were largely replicated in ABIDE2 (Figure 1A, Supplementary Table S3-S4). The unthresholded ABIDE1 and ABIDE2 LCOR t-contrast maps displayed a strong positive spatial association (*r* = .597, *p* < .001) indicating a similar topology of ASD alterations across both cohorts.

**Fig. 1.**
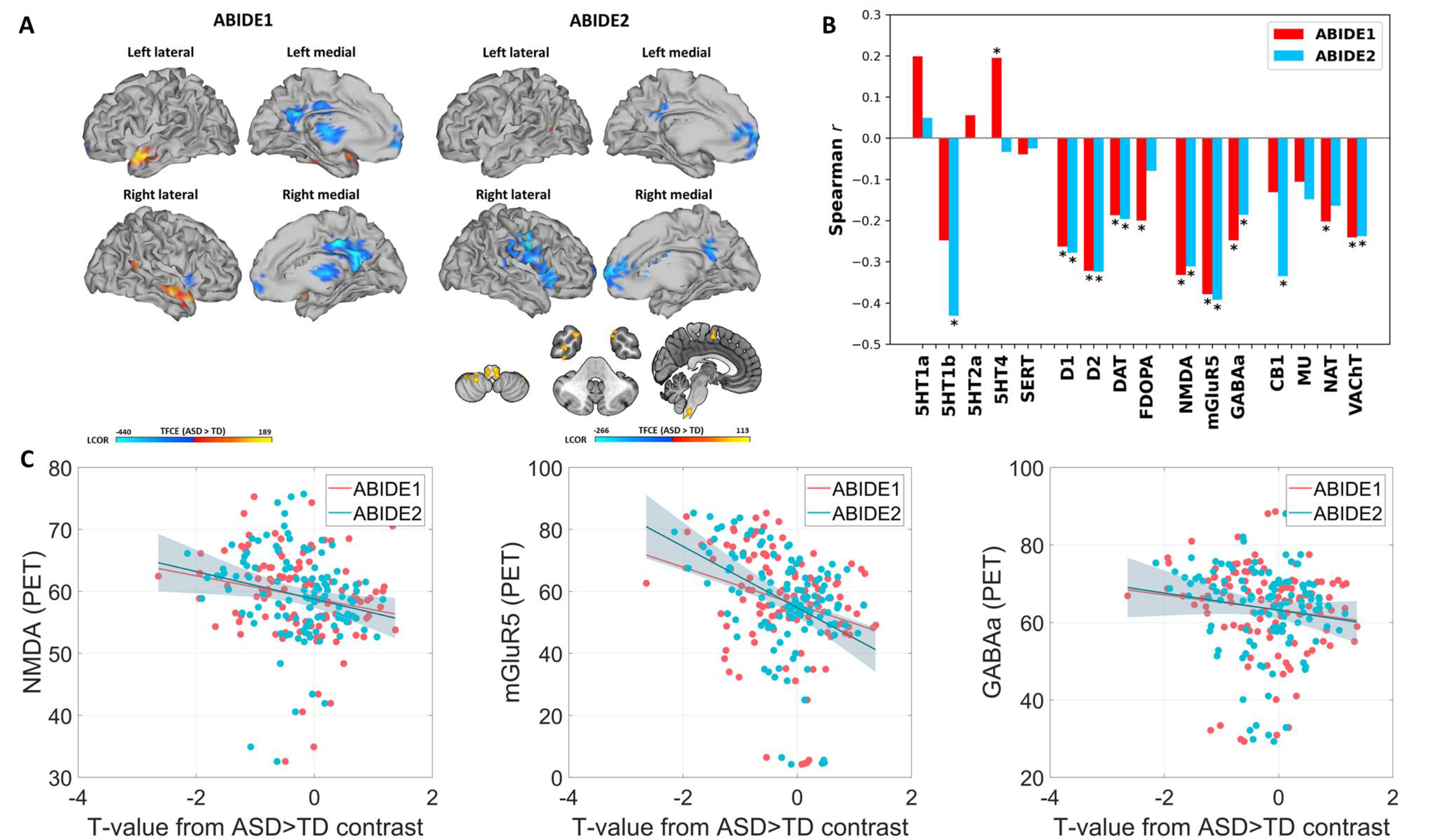
Local functional activity alterations in subjects with autism spectrum disorder (ASD) and its neurotransmitter co-localizations. (A) Results from voxel-wise comparisons between subjects with ASD and typically developed controls (TD) regarding local synchronization (LCOR). Positive T-values (red-yellow) indicate increased LCOR in ASD subjects compared to TD, negative T-values (blue) represent decreased LCOR. Left: Results from voxel-wise comparisons within the Autism Brain Imaging Data Exchange (ABIDE) 1 (sagittal view). Right: Results from voxel-wise comparisons within the replication dataset ABIDE2 (sagittal and axial view). (B) Grouped bar plot depicting the spatial co-localizations of whole-brain LCOR alterations in subjects with ASD compared to TD with 16 different receptor and transporter distributions. The asterisk (*) represents significant co-localizations (*p* < .05). (C) Scatterplots showing the negative relationship between LCOR alterations in ASD (T-values) and the nuclear imaging derived spatial NMDA, mGluR5 and GABAa receptor distributions.

### LCOR alterations in ASD co-localize with specific neurotransmitter systems

Next, we evaluated whether LCOR alterations observed in ASD were spatially linked to the distribution of specific neurotransmitter systems derived from nuclear imaging in healthy volunteers. Whole-brain LCOR alterations in ASD subjects in ABIDE1 were found to significantly co-localize with the spatial distribution of dopaminergic D1 (*r* = -.263, *p* = .005) and D2 receptors (*r* = -.322, *p* < .001) as well as DAT (*r* = -.187, *p* = .033) and FDOPA (*r* = -.200, *p* = .034). Further significant associations were observed with serotonergic 5HT4 receptors (*r* = .195, *p* = .024), the ionotropic NMDA receptor (*r* = -.332, *p* = .009), the metabotropic mGluR5 receptors (*r* = -.379, *p* = .004), GABAa receptors (*r* = -.248, *p* = .005), NAT (*r* = -.202, *p* = .024) and the cholinergic transporter VAChT (*r* = -.241, *p* = .003). The significant co-localizations with D1, D2, DAT, NMDA, mGluR5, GABAa and VAChT were replicated in the ABIDE2 dataset (Figure 1B-C, Supplementary Table S5).

### Ketamine- and midazolam-induced LCOR changes co-localize with neurotransmitter systems

Similarly to the effects of ASD, we tested for co-localization of ketamine- and midazolam-induced changes (compared to placebo) with specific neurotransmitter systems. Ketamine administration induced LCOR changes significantly co-localized with D1 (*r* = -.282, *p-FDR* = .032), NMDA (*r* = -.338, *p-FDR* = .032) and GABAa receptors (*r* = -.251, *p-FDR* = .037) (Supplementary Table S6 and Supplementary Figure S1). The effect of midazolam on LCOR co- localized with the distribution of 5HT2a (*r* = -.305, *p-FDR* = .047), SERT (*r* = .436, *p-FDR* < .001), DAT (*r* = .426, *p-FDR* = .002), FDOPA (*r* = .417, *p-FDR* < .001), CB1 (*r* = -.468, *p-FDR* = .002), NAT (*r* = .264, *p-FDR* = .019) and VAChT (*r* = .332, *p-FDR* < .001).

### ASD-related alterations co-localize with ketamine effects

We tested for similarity of ASD-related LCOR brain patterns with alterations induced by NMDA-receptor-antagonist ketamine and GABAa-potentiator midazolam (s. Supplementary Material S7-10 and Supplementary Figure S2-3 for detailed description and visualization of LCOR changes induced by ketamine and midazolam compared to placebo). Whole-brain LCOR alterations observed across both ABIDE datasets showed a significant positive correlation with ketamine-induced changes (ABIDE1: *r* = .381, *p* = .003; ABIDE2: *r* = .292, *p* = .036). The effects of midazolam displayed no significant association with ASD-related LCOR alterations (ABIDE1: *r* = .018, *p* = .903; ABIDE2: *r* = .205, *p* = .155). The ketamine and midazolam whole-brain *t*- contrast maps did not show a significant association (Supplementary Table S11).

We then tested for ketamine- and midazolam-induced LCOR changes in regions significantly altered in ASD by computing *t*-tests for ΔKET and ΔMDZ changes (vs. placebo).

Ketamine significantly reduced LCOR in regions showing a reduction in ASD (Figure 2A) (T = - 1.978, *p* = .029, Cohen’s d = 0.299). Midazolam administration increased LCOR in regions displaying an increase in ASD (T = 2.011, *p* = .027, Cohen’s d = .270).

**Fig. 2.**
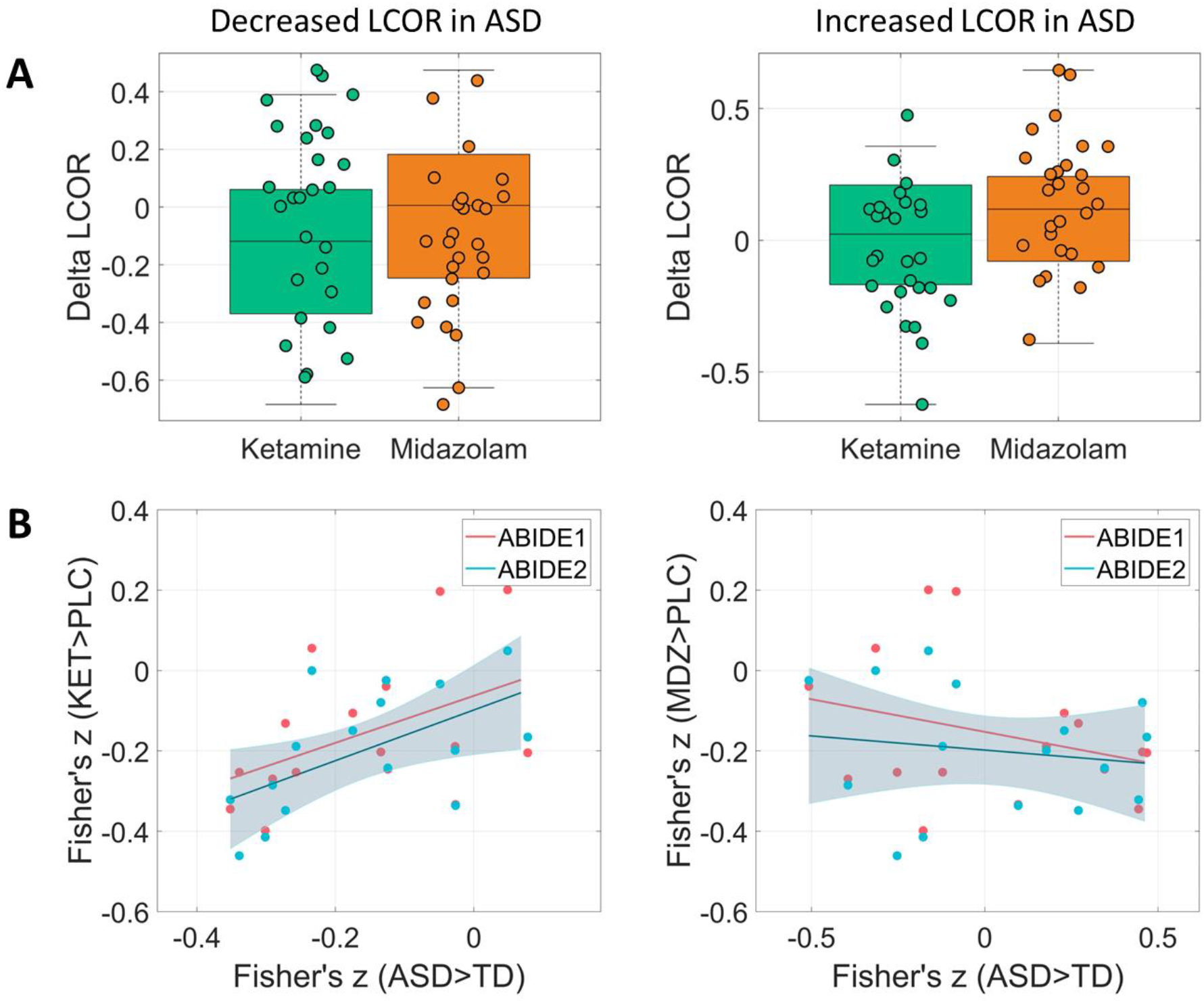
Mean local activity (LCOR) after ketamine (KET) and midazolam (MDZ) administration in regions significantly altered in autism spectrum disorder (ASD) and correlation of the neurochemical co-localization profiles between ASD, KET and MDZ condition. (A) Boxplots comparing LCOR following KET and MDZ administration relative to placebo (PLC) condition. Delta LCOR is defined as the mean LCOR in the KET or MDZ condition minus the mean LCOR in the PLC condition. The plots display Delta LCOR values within all voxels that showed significant decreases (left) or increases (right) in individuals with ASD compared to typically developing controls (TD) in the Autism Brain Imaging Data Exchange 1 (ABIDE1) dataset. (B) Scatterplots illustrating the relationship between the 16 Fisher’s z-values for KET (left) and MDZ (right) versus PLC, and ASD versus TD in the ABIDE1 and ABIDE2 datasets. The 16 Fisher’s z-values represent the LCOR-neurotransmitter co-localization profiles for the respective contrast.

### Neurochemical co-localization profiles are similar between ASD and ketamine conditions

Next, we tested for the overall similarity of the pharmacological neurotransmitter co- localization profiles with those observed in individuals with ASD across all evaluated neurotransmitter systems. For this, we extracted mean Fisher’s z values for the 16 tested receptors and transporters for ASD, ketamine and midazolam-related changes and computed Pearson correlations between these profiles. We found a significant association between the neurochemical profiles observed for ketamine and ASD in ABIDE2 (*r* = .563, *p* = .023) and a trend towards significance for ABIDE1 (*r* = .448, *p* = .081) (Figure 2B). The neurochemical profile induced by midazolam did not show a significant association with the ASD profile (ABIDE1: *r* = -.241, *p* = .368; ABIDE2: *r* = .173, *p* = .521). Notably, the neurochemical ASD profiles across ABIDE1 and ABIDE2 datasets were highly correlated (*r* = .814, *p* < .001). The neurochemical profiles for ketamine and midazolam were not significantly correlated (*r* = .407, *p* = .118) (Supplementary Table S6).

### No consistent association of the LCOR-neurotransmitter co-localizations in ASD with clinical symptom domains

Lastly, we tested for associations between individual z-scores of ASD subjects for the replicated whole-brain LCOR-neurotransmitter co-localization findings and different ASD symptom domains as measured with the Autism Diagnostic Observation Schedule (ADOS)^32^ (Supplementary Table S12). In ABIDE1, no significant associations were found with the ADOS total score nor the three subscores (communication, social interaction, stereotyped behavior and restricted interests [SBRI]). A significant but weak negative association was observed in ABIDE2 between SBRI and the strength of LCOR-GABAa co-localizations (*r* = -.133, *p* = .040) and a positive with the strength of LCOR-VAChT co-localizations (*r* = .141, *p* = .030) (Supplementary Fig. S4). Both findings did not survive a correction for multiple comparisons.

## DISCUSSION

Here, we provide evidence for consistent local functional activity (LCOR) decreases in individuals with ASD. These alterations co-localized with *in-vivo* derived distributions of specific receptors and transporters covering dopaminergic, glutamatergic, GABAergic and cholinergic neurotransmission. The LCOR pattern observed in ASD across both cohorts was similar to the effect of NMDA-antagonist ketamine but not to the effect of the GABAergic medication midazolam. Ketamine reduced LCOR in regions exhibiting decreases in ASD. At the neurochemical level, the LCOR-neurotransmitter co-localization profile of ketamine was similar to changes observed in ASD.

Previous studies investigating functional alterations in ASD have often yielded inconsistent findings, potentially due to small sample sizes, the phenotypic heterogeneity of ASD, or varying methodological approaches^4^. Our results support the notion of consistent group-level local activity decreases as measured using LCOR in brain regions implicated in self-referential processing, social cognition, cognitive and emotional regulation^33–36^. These findings align with recent rs-fMRI meta-analyses indicating local hypoactivity within the PCC^6^, precuneus and right temporal gyri^9^ and reinforce the role of functional abnormalities of the DMN in the pathophysiology of ASD^8,11^. Other studies utilizing ABIDE or other datasets have reported comparable alterations in regional homogeneity (ReHo), a metric closely related to LCOR^37,38^. Additionally, task-based fMRI studies have demonstrated reduced activation in these regions in ASD during tasks that require self-related versus other-related judgements^39,40^. This suggests that decreased LCOR in DMN nodes may relate to difficulties of individuals with ASD in self-referential processing and inferring mental states of others (“theory of mind”). Our findings further support recent efforts to identify neuro-subtypes in ASD, which have observed converging abnormalities in both the DMN and frontoparietal networks across subgroups^41,42^. Conversely, we did not observe consistent LCOR increases across both ABIDE datasets.

Most previous studies were restricted to the investigation of macroscale brain networks based on hemodynamic signals of rs-fMRI. To gain a deeper understanding of the neurobiological mechanisms underlying the emergent functional activity alterations in ASD, it is crucial to integrate data from molecular neuroimaging. This approach could extend the brain connectivity framework by incorporating a molecular perspective^12,14,30^. Adopting this approach, we found LCOR alterations observed across both ABIDE datasets to be spatially related to dopaminergic, glutamatergic, GABAergic and cholinergic neurotransmission. Specifically, all co-localizations were negative with stronger LCOR reductions in ASD being associated with increased availability of respective receptors and transporters in health. In line with the observed clinical heterogeneity and recently suggested existence of ASD subtypes^41^, these findings support the notion of involvement of multiple neurotransmitter systems in the pathogenesis of ASD.

We observed the strongest similarity of ASD patterns with glutamatergic NMDA and mGluR5 receptor distributions, which represents the main excitatory neurotransmitter system. Glutamate acts in a homeostatic relationship with the inhibitory GABA system balancing neuronal excitability^43^. The glutamatergic system plays an important role in brain development and neuroplasticity^13^. A balanced E/I ratio is considered to be crucial for maintaining mental health, with better overall cognitive performance in healthy children associated with a lower E/I ratio, particularly in higher-order association cortices such as the DMN nodes^44^. Conversely, an increased E/I ratio has been frequently implicated in the ASD phenotype^25,26^. For instance, elevated glutamate levels, which can be found in ASD in peripheral blood serum and within the central nervous system^19,45^, have been shown to have neurotoxic effects potentially leading to neuronal cell death and volumetric reductions as shown in structural MRI studies^46^. A previous multimodal imaging study observed a negative association between increased glutamate concentrations in the dorsal ACC and reduced functional connectivity between the dorsal ACC and insular, limbic and parietal regions in male subjects with ASD^22^. The dorsal ACC plays an important role in top-down cognitive control and adaptive, flexible behavior with decreased functional connectivity in this region linked to deficient response inhibition and more repetitive behavior in ASD^33^. Consistently, higher mGluR5 densities can be observed in male adults with ASD^20^.

The crucial role of glutamate dysregulation in the pathogenesis of ASD has been highlighted by pharmacological studies manipulating the glutamatergic system^21^. Although the results are inconsistent, D-Cycloserine, a NMDA receptor agonist, showed positive effects in treatment of social impairment and stereotypies in subjects with ASD^47,48^. Conversely, the uncompetitive NMDA receptor antagonist ketamine led to reduced mentalizing performance in a social cognition task associated with increased neural activity in the superior temporal sulcus and anterior precuneus as well as increased psychotic symptoms in healthy adults^29^. In accordance, early postnatal ketamine administration in mice increased stereotyped behavior and social impairment in later adulthood combined with elevated glutamate and reduced GABA levels in the amygdala and hippocampus^49^. In clinical studies, intranasal ketamine administration in adolescents and young adults with ASD showed no improvement of ASD core symptoms^50^. Conceptually, one would expect a successful treatment to normalize brain functional patterns associated with a specific clinical condition. Our own findings are therefore in line with the negative outcome of the ketamine trial, showing that ketamine actually induces ASD-like brain patterns by reducing LCOR in regions where it is also reduced in ASD. Corroborating this finding, the neurochemical profile induced by ketamine significantly correlates with the neurochemical profile found in ASD across all tested neurotransmitter systems. Specifically, the effect of ketamine on LCOR co-localized with the distribution of NMDA, mGluR5 and GABAa receptors suggesting that ketamine may induce ASD-like activity patterns through disturbing the E/I balance. Future pharmacological studies should investigate the effectiveness of drugs manipulating this balance opposing the effects of ketamine in relieving ASD symptoms.

Optogenetic studies demonstrated that experimentally elevating the E/I ratio within the rodent medial prefrontal cortex, an important DMN hub, caused impaired information processing and social dysfunction^51^. Notably, compensatory elevation of inhibitory neurons alleviated social dysfunction by balancing the E/I ratio. Potentially, an atypical expression of GABAa within the DMN contributes to ASD symptom severity^23,52^. For instance, Oblak et al. (2010) found reduced GABAa receptors and benzodiazepine binding sites in the PCC and fusiform gyrus post mortem in subjects with ASD^53^. Additionally, meta-analyses accumulate evidence for a negative association between local GABA concentrations and functional activation of the medial prefrontal cortex and ACC during emotion processing tasks^54^. Within our study, pharmacologically increased inhibition by GABAa-potentiator midazolam in healthy volunteers did not affect LCOR in the DMN regions where LCOR was decreased in ASD. Conversely, midazolam significantly elevated LCOR in regions where LCOR was increased in ASD, indicating a partial induction of ASD-like brain patterns and complementing the findings observed with ketamine. At the neurochemical level, midazolam effects did not co-localize with the neurochemical signature of ASD (Figure 2B, Supplementary Figure 5). To date, there is modest evidence of the effectiveness of GABA modulators such as arbaclofen and acamprosate in treating ASD symptoms^28,55^.

Several studies support the notion of altered dopamine and serotonin neurotransmission in ASD, a hypothesis further reinforced by clinical observations indicating a small positive effect of the atypical antipsychotics aripiprazole and risperidone, as well as selective serotonin reuptake inhibitors, on alleviating stereotypies and aggressive behavior in some ASD subgroups^13,17,56^. For instance, an overexpression of the dopaminergic system has been implicated in the modulation of stereotyped behavior in ASD animal models^15^. The dopamine hypothesis of ASD suggests that social deficits may result from dysfunction in the mesocorticolimbic system, while stereotypies are thought to arise from abnormalities within the nigrostriatal circuitry^16^. We observed that LCOR reductions in ASD were negatively co-localized with dopaminergic D1/D2 receptors and the dopamine transporter. These co-localizations did not yield significant correlations with specific symptom domains. Similarly, we did not find consistent *in-vivo* co-localization between LCOR and serotonergic neurotransmission across the ABIDE cohorts. Elevated blood serotonin levels have been reported in only a subset of autistic children (approximately 25%), and much of the supporting evidence comes from animal models^17^. For both of these analyses, our restriction to whole-brain analyses and disregard of potential subtypes may have limited the ability to detect some regionally and subtype specific co-localization patterns.

In that regard it is also important to note that the included multi-site datasets are extremely heterogeneous with respect to availability of demographic, medication status and clinical information as well as in terms of image quality. This variability may obscure more nuanced insights into the neurobiological mechanisms underlying ASD. Future research should aim to conduct large-scale standardized studies to allow for identification of distinct ASD subtypes and their associated neurobiological profiles.

## Conclusions

We find LCOR alterations in subjects with ASD to negatively co-localize with the *in-vivo* derived distribution of dopaminergic, glutamatergic, GABAergic and cholinergic neurotransmitter systems in two large independent cohorts. This pattern of LCOR alterations in ASD was similar to the effect induced by the NMDA-antagonist ketamine. These findings provide novel insights into the pathophysiology of ASD-related functional alterations and may guide new hypotheses for pharmacological interventions. Future neuro-subtyping studies aimed at disentangling the heterogeneity within ASD will be crucial for identifying biologically and clinically distinct subgroups that allow for precision psychiatry approaches in ASD.

## MATERIALS AND METHODS

### Study design

We included data from the open-access Autism Brain Imaging Data Exchange 1 (ABIDE1) into the study, which involves 1,112 participants (ASD: N = 539) from 17 international centers sharing rs-fMRI and corresponding anatomical T1-weighted images as well as phenotypic data^57^. We additionally included the independently collected ABIDE2 dataset for subsequent replication of any results observed in the ABIDE1 cohort^58^. ABIDE2 includes data from 1,114 participants (ASD: N = 521) from 19 centers. For further information of the varying recruitment and diagnostic criteria, please see https://fcon_1000.projects.nitrc.org/indi/abide. We excluded subjects with intellectual disability (IQ ≤ 70) to reduce variability associated with low-functioning ASD, subjects with missing data and subjects with excessive head motion during image acquisition (translation > 3 mm or rotation > 3°). For ABIDE2, another 3 subjects were excluded due to preprocessing failure of imaging data (Supplementary Fig. S5). Thus, 878 subjects (405 with ASD) from ABIDE1 and 869 subjects (395 with ASD) from the ABIDE2 dataset were included in the analyses (Table 1).

For the comparison between pharmacological brain signatures and aberrant local functional activity in ASD, we used data from 30 healthy male participants without a history of recreational drug use, who were scanned in a single-blinded, placebo-controlled, three-way cross-over design receiving ketamine, midazolam or placebo. The substances were administered to a sub-anesthetic level through intravenous access by an infusion pump. This cohort and the assessment procedures have been extensively described in Forsyth et al. (2020)^59^. Three participants were excluded following image quality control, so that the final dataset consisted of 27 participants (M = 26.6 ± 5.9 years, 19-37 years).

All participants gave written informed consent, and all studies were approved by local ethics committees.

### MRI data acquisition

As ABIDE1 and 2 data was retrospectively collected from multiple centers, acquisition of MRI images varied between sites. For individual site profiles regarding MRI protocol information, see Supplementary Table S13-14.

MRI images of the pharmacological dataset were acquired on a 3T MRI scanner (Siemens Skyra, Erlangen, Germany) with a 20-channel head coil. For structural imaging, a 3D magnetization-prepared rapid gradient-echo (3D-MPRAGE) scan [echo time (TE) = 3.42 ms; repetition time (TR) = 2100 ms; FOV = 256 mm^2^; flip angle 9°; 192 slices; slice thickness = 2 mm; voxel size = 1x1x1 mm] was acquired. Additionally, 246 volumes of BOLD rs-fMRI data were obtained using a T2*-weighted echo planar imaging (EPI) sequence (TE = 27 ms; TR = 2200 ms; flip angle 79°; 30 interleaved 3 mm slices; voxel size = 3x3x3 mm). During imaging, participants wore an EEG-cap, which was removed in one of the three sessions for high resolution 3D-MPRAGE structural image acquisition^59^.

### Preprocessing of imaging data

All rs-fMRI and structural imaging data were preprocessed using Statistical Parametric Mapping software SPM12^60^ and the CONN toolbox^61^ implemented in Matlab (v2022b). Functional images were corrected for head motion and distortions (realign and unwarp), spatially normalized into MNI space and resampled to a resolution of 2 mm^3^ isotropic. Smoothing was applied using a 6 mm full-width at half maximum (FWHM) Gaussian kernel. Mean white matter, grey matter and cerebrospinal fluid signals, as well as 24 motion parameters were regressed out before computing the voxel-based measures using the CONN toolbox^62^. Motion parameters were used to identify data to be excluded due to excessive head movement (translation > 3 mm and rotation > 3°). For the pharmaco-fMRI dataset, preprocessing was applied equivalently to the three pharmacological conditions (ketamine, midazolam and placebo).

### Resting-state functional activity measure

To assess functional activity, we examined local synchronization (LCOR) by computing correlation at each voxel. LCOR was chosen as it provides a good approximation of local metabolism and has been recently shown to be sensitive to local pathological functional changes across different neurotransmitter systems^63,64^. LCOR is defined as the average correlation between a given voxel with other voxels in its proximity, with distances weighted by a Gaussian kernel (25 mm FWHM)^65^.

### Voxel-wise group comparisons

To test for LCOR changes in ASD, we performed group comparisons between subjects with ASD and TD controls using the CONN toolbox. We utilized the ABIDE1 dataset for initial exploration and validated our findings using the ABIDE2 dataset. All group comparisons were corrected for age, sex, full scale IQ (FIQ), motion and site. Pairwise *t*-contrasts comparing ASD and TD were evaluated for significance using a voxel-wise family-wise error threshold of *p* < 0.05 combined with an exact permutation-based cluster threshold (threshold free cluster enhancement (TFCE), 1000 permutations, *p* < 0.05) to control for multiple testing. The same procedure was applied to the pharmaco-fMRI dataset comparing the respective pharmacological condition with placebo condition.

### Co-localization between LCOR and neurotransmitter maps

The JuSpace toolbox was used for all further co-localization analyses^66^. Distribution of neurotransmitter systems were derived from positron emission and single photon emission computer tomography (PET, SPECT) from independent healthy volunteer populations. Neurotransmitter maps were available for serotonergic (5-HT1a, 5-HT1b, 5-HT2a, 5HT4) and dopaminergic (D1, D2) receptors, the dopamine (DAT) and serotonin transporter (SERT), the dopamine synthesis capacity (fluorodopa PET - FDOPA), GABAa, ionotropic NMDA and metabotropic glutamate receptor (mGluR5), μ-opioid (MU) and cannabinoid receptors (CB1), noradrenaline transporter (NAT) and vesicular acetylcholine transporter (VAChT).

We first examined whether ASD related alterations in LCOR were spatially similar to the above neurotransmitter systems correlating the respective PET maps with the unthresholded t-contrast maps of LCOR alterations observed in ASD (from ASD>TD contrast). For this, we adopted an exploration and replication approach. We first tested for significant co-localizations in ABIDE1 applying an uncorrected p-value of *p* < 0.05 (two-sided) and then tested for replication of the significant findings in ABIDE2. The default Neuromorphometrics atlas (119 regions) covering cortical, subcortical and cerebellar regions of interest was used for all analyses. Partial Spearman correlations were computed adjusting for spatial autocorrelation using the gray matter probability map. Exact permutation-based p-values were computed comparing the observed correlation coefficients to those obtained using permuted PET maps whilst preserving the spatial autocorrelation. A detailed description of the workflow within the JuSpace toolbox is provided by Dukart et al. (2021)^66^.

This procedure was repeated with the pharmacological dataset testing for co-localization of the unthresholded t-contrasts for LCOR changes induced by ketamine and midazolam (both testing for increases over placebo) with the respective neurotransmitter maps. The Benjamini-Hochberg procedure was used to account for multiple comparisons.

### Correlation of ASD-related LCOR alterations with ketamine- and midazolam-induced brain changes

To test for similarity between functional alterations induced by NMDA-antagonist ketamine and GABAa-potentiator midazolam and ASD-related alterations, we first spatially correlated the respective whole-brain LCOR t-contrast maps. To assess the impact of glutamatergic and GABAergic medication on brain regions exhibiting significant differences between ASD and TD groups, we separately extracted mean LCOR values per subject for ketamine, midazolam and placebo conditions from regions showing significant increases or decreases in ASD in the ABIDE1 dataset. We then calculated Δ-scores for each subject in regions showing either increases or decreases in ASD (separately) by subtracting LCOR values for placebo from ketamine and midazolam conditions. We then performed one-sample *t*-tests for ΔKET and ΔMDZ based on the hypothesis that both drugs induce ASD-like brain patterns (*p* < 0.05 one-sided).

Lastly, we tested for similarity of the mappings of ASD alterations onto all neurotransmitter systems with those observed for ketamine and midazolam. For this, we first extracted the 16 Fisher’s z-transformed Spearman correlations observed for the co-localization between ASD-related alterations and all of the above neurotransmitter maps. Similarly, we extracted the correlation values observed for the co-localization of changes induced by ketamine and midazolam with the same 16 neurotransmitter maps. We computed Pearson correlations between these neurochemical co-localization profiles obtained for ASD, and ketamine and midazolam conditions.

### Relationship of LCOR-neurotransmitter co-localizations with clinical phenotypes of ASD

Next, we tested whether the significant co-localizations of LCOR with specific neurotransmitters across both ABIDE cohorts relate to symptom severity in ASD. For this, we calculated individual z-scores in each region for each ASD subject relative to the mean of the respective TD control group and correlated the obtained z-maps with the respective neurotransmitter maps. The obtained individual Fisher’s z-transformed coefficients were used to perform Pearson correlations with clinical measures of autistic symptom severity within IBM SPSS Statistics (Version 27)^67^.

We used the Autism Diagnostic Observation Schedule (ADOS) as a measure of ASD symptom severity^32^. The ADOS is a semi structured, standardized measure of autistic phenotypes which consists of four age-adjusted 30-minute modules according to the level of expressive language covering the following three subdomains: communication deficits, reciprocal social interaction deficits (RSI), and stereotyped behaviors and restricted interest (SBRI) (Table 1).

## Data Availability

Ketamine and midazolam data are available upon reasonable request to the authors.
The ABIDE datasets are available online at https://fcon_1000.projects.nitrc.org/indi/abide/

## List of Supplementary Materials

### Results

Table S1-S4. Local functional activity alterations in ASD compared to TD.

Table S5. LCOR alterations in ASD relate to the *in-vivo* distribution of neurotransmitter systems.

Table S6. Neurotransmitter co-localizations with LCOR changes induced by ketamine and midazolam.

Fig. S1. Visualization of ketamine and midazolam co-localization profiles.

Table S7-10. LCOR alterations induced by ketamine and midazolam administration.

Fig. S2-3. Visualization of ketamine and midazolam induced LCOR changes.

Table S11. Similarity of ASD and medication effects.

Table S12. Correlation of the LCOR-neurotransmitter co-localizations with ASD symptom domains in ABIDE1 and ABIDE2.

Fig. S4. Correlation of the LCOR-neurotransmitter co-localizations in ASD with clinical symptom domains.

### Materials and Methods

Fig. S5. Exclusion process.

Table S13-14. MRI data acquisition.

## Acknowledgments

We thank all participants for taking part in this research. JD has received funding from the European Union’s Horizon 2020 research and innovation program under grant agreement No. 826421, “TheVirtualBrain-Cloud.” SBE has received funding from the European Union’s Horizon 2020 research and innovation program under grant agreement No. 945539 (Human Brain Project SGA3). ABIDE I is supported by NIMH (K23MH087770), NIMH (R03MH096321), Leon Levy

Foundation, Joseph P. Healy, and the Stavros Niarchos Foundation. ABIDE II is supported by NIMH (5R21MH107045), NIMH (5R21MH107045), Nathan S. Kline Institute of Psychiatric Research, Joseph P. Healey, Phyllis Green, and Randolph Cowen.

## Author contributions

Conceptualization: PG, JD

Methodology: PG, JD

Formal analysis: PG, JD

Investigation: PG, JD

Data curation: JFH, AF, SM, JD

Visualization: PG, JK, JD

Project administration: JD

Supervision: JD

Writing – original draft: PG

Writing – review & editing: JK, JFH, AF, SLV, SM, SBE, LS, JD

## Data and materials availability

All data are available in the main text or the supplementary materials.

