## Supplementary Material for "Local activity alterations in autism spectrum disorder correlate with neurotransmitter properties and ketamine induced brain changes"

**Results**

**Supplementary Table S1-4. Local functional activity alterations in ASD compared to TD**

| **contrast** | **cluster (x, y, z)** | **size** | **peaks** | **TFCE** | **peak *p-FWE*** | **peak *p-FDR*** |
| --- | --- | --- | --- | --- | --- | --- |
| **ASD>TD** | -45 +00 -24 | 67 | 4 | 313.04 | .0030 | .0044 |
|  | +51 -12 -18 | 87 | 7 | 220.45 | .0100 | .0070 |
|  | -33 -21 -15 | 12 | 2 | 201.43 | .0150 | .0070 |
|  | +51 -54 +12 | 14 | 1 | 185.57 | .0220 | .0070 |
|  | -42 -36 -30 | 10 | 1 | 182.86 | .0300 | .0070 |
|  | -33 +03 -45 | 14 | 4 | 177.16 | .0350 | .0070 |
|  | -27 -15 -27 | 6 | 1 | 175.80 | .0390 | .0070 |
|  | +27 +06 -39 | 3 | 1 | 175.59 | .0400 | .0070 |
|  | +48 -63 +18 | 6 | 1 | 173.68 | .0420 | .0070 |
|  | +60 -48 +09 | 5 | 1 | 172.69 | .0430 | .0070 |
|  | -36 -12 -27 | 2 | 1 | 169.24 | .0440 | .0070 |
|  | +57 -39 +00 | 5 | 1 | 168.51 | .0440 | .0070 |
|  | -45 -30 -30 | 5 | 1 | 167.35 | .0460 | .0070 |
|  | -51 -33 -03 | 7 | 1 | 166.27 | .0460 | .0070 |
|  | -51-39 +06 | 3 | 1 | 166.06 | .0460 | .0070 |
|  | -36 -39 -21 | 1 | 1 | 165.07 | .0470 | .0070 |
|  | -30 -03 -45 | 1 | 1 | 163.23 | .0490 | .0070 |
| **ASD<TD** | -03 -51 +21 | 491 | 15 | -596.38 | < .0001 | < .0001 |
|  | +00 -21 +06 | 151 | 7 | -368.78 | < .0001 | < .0001 |
|  | +45 +09 -06 | 47 | 2 | -236.67 | .0040 | .0007 |
|  | -09 +54 -09 | 72 | 7 | -225.19 | .0050 | .0011 |
|  | +00 +42 -06 | 29 | 3 | -190.17 | .0170 | .0027 |
|  | -06 +57 +09 | 23 | 3 | -185.78 | .0220 | .0029 |
|  | -03 +12 +27 | 15 | 2 | -170.32 | .0340 | .0040 |
| **Supplementary Table S1.** Voxel-wise comparisons between ASD and TD regarding LCOR in the ABIDE1 cohort. List of significant TFCE-corrected cluster for positive (ASD>TD) and negative contrast (ASD<TD); highlighted cluster coordinates indicate the voxel with the highest T-value within a cluster; ABIDE = autism brain imaging data exchange; ASD = autism spectrum disorder; TD = typically developed controls; TFCE = threshold free cluster enhancement; FDR = false discovery rate; FWE = family-wise error; LCOR = local synchronization; TFCE-threshold: *p* < 0.05. | | | | | | |

We found a replicable pattern of local synchronization (LCOR) reductions in individuals with autism spectrum disorder (ASD) in the default mode network (DMN), anterior cingulate cortex, paracingulate gyrus, precentral gyrus and right insular and opercular cortex. Increased LCOR in ASD was found in bilateral temporal regions, the cerebellum, right angular gyrus and lateral occipital cortex.

Contrast: ASD>TD

28 voxels (11%) covering 4% of atlas.TP l (Temporal Pole Left)

23 voxels (9%) covering 18% of atlas.aMTG l (Middle Temporal Gyrus, anterior division Left)

22 voxels (9%) covering 3% of atlas.TP r (Temporal Pole Right)

19 voxels (8%) covering 23% of atlas.aSTG r (Superior Temporal Gyrus, anterior division Right)

13 voxels (5%) covering 10% of atlas.aMTG r (Middle Temporal Gyrus, anterior division Right)

13 voxels (5%) covering 3% of atlas.pMTG r (Middle Temporal Gyrus, posterior division Right)

12 voxels (5%) covering 5% of atlas.Hippocampus l

11 voxels (4%) covering 4% of atlas.pTFusC l (Temporal Fusiform Cortex, posterior division Left)

8 voxels (3%) covering 2% of atlas.toMTG r (Middle Temporal Gyrus, temporooccipital part Right)

7 voxels (3%) covering 8% of atlas.aSTG l (Superior Temporal Gyrus, anterior division Left)

7 voxels (3%) covering 2% of atlas.pMTG l (Middle Temporal Gyrus, posterior division Left)

6 voxels (2%) covering 0% of atlas.sLOC r (Lateral Occipital Cortex, superior division Right)

6 voxels (2%) covering 6% of atlas.aTFusC l (Temporal Fusiform Cortex, anterior division Left)

5 voxels (2%) covering 4% of atlas.pSTG l (Superior Temporal Gyrus, posterior division Left)

5 voxels (2%) covering 1% of atlas.AG r (Angular Gyrus Right)

4 voxels (2%) covering 1% of atlas.pITG l (Inferior Temporal Gyrus, posterior division Left)

2 voxels (1%) covering 2% of atlas.pSTG r (Superior Temporal Gyrus, posterior division Right)

1 voxels (0%) covering 1% of atlas.aITG l (Inferior Temporal Gyrus, anterior division Left)

1 voxels (0%) covering 1% of atlas.aPaHC r (Parahippocampal Gyrus, anterior division Right)

1 voxels (0%) covering 1% of atlas.PP r (Planum Polare Right)

1 voxels (0%) covering 0% of atlas.Cereb6 l (Cerebelum 6 Left)

53 voxels (21%) covering 0% of atlas.not-labeled

Contrast: ASD<TD

323 voxels (39%) covering 45% of atlas.PC (Cingulate Gyrus, posterior division)

136 voxels (16%) covering 8% of atlas.Precuneous (Precuneous Cortex)

83 voxels (10%) covering 20% of atlas.Thalamus l

63 voxels (8%) covering 16% of atlas.Thalamus r

31 voxels (4%) covering 8% of atlas.IC r (Insular Cortex Right)

29 voxels (4%) covering 1% of atlas.FP l (Frontal Pole Left)

28 voxels (3%) covering 4% of atlas.AC (Cingulate Gyrus, anterior division)

24 voxels (3%) covering 1% of atlas.FP r (Frontal Pole Right)

21 voxels (3%) covering 6% of atlas.PaCiG l (Paracingulate Gyrus Left)

19 voxels (2%) covering 7% of atlas.MedFC (Frontal Medial Cortex)

9 voxels (1%) covering 2% of atlas.PaCiG r (Paracingulate Gyrus Right)

5 voxels (1%) covering 1% of atlas.TP r (Temporal Pole Right)

3 voxels (0%) covering 0% of atlas.PreCG l (Precentral Gyrus Left)

2 voxels (0%) covering 1% of atlas.IFG oper r (Inferior Frontal Gyrus, pars opercularis Right)

1 voxels (0%) covering 0% of atlas.PreCG r (Precentral Gyrus Right)

1 voxels (0%) covering 1% of atlas.FO r (Frontal Operculum Cortex Right)

1 voxels (0%) covering 0% of atlas.CO r (Central Opercular Cortex Right)

49 voxels (6%) covering 0% of atlas.not-labeled

**Supplementary Table S2.** Regions with significant voxels in ABIDE1 (all clusters combined). List of all significant voxels and atlas regions in the autism brain imaging data exchange (ABIDE) 1 dataset for the positive (ASD>TD) and negative contrast (ASD<TD); ASD = autism spectrum disorder; TD = typically developed controls.

| **contrast** | **cluster (x, y, z)** | **size** | **peaks** | **TFCE** | **peak *p-FWE*** | **peak *p-FDR*** |
| --- | --- | --- | --- | --- | --- | --- |
| **ASD>TD** | -06 -39 -60 | 46 | 4 | -186.59 | .0220 | .0157 |
|  | -39 -63 +09 | 10 | 1 | -183.92 | .0240 | .0157 |
|  | +06 -03 +60 | 13 | 2 | -176.54 | .0280 | .0157 |
|  | +18 -48 -57 | 11 | 3 | -172.64 | .0320 | .0157 |
|  | -21 +15 -42 | 7 | 2 | -171.96 | .0320 | .0157 |
|  | -18 -39 -54 | 4 | 1 | -167.94 | .0370 | .0157 |
|  | +30 +12 -42 | 14 | 1 | -166.09 | .0390 | .0157 |
|  | -36 -57 -60 | 6 | 1 | -165.65 | .0400 | .0157 |
|  | -21 +06 -45 | 5 | 2 | -163.34 | .0410 | .0157 |
|  | +42 +06 -45 | 5 | 1 | -160.92 | .0460 | .0157 |
|  | +30 -48 -57 | 4 | 1 | -160.61 | .0470 | .0157 |
|  | +45 -48 +45 | 4 | 1 | -160.45 | .0470 | .0157 |
|  | +39 -06 -42 | 8 | 1 | -159.69 | .0470 | .0157 |
| **ASD<TD** | -03 +48 +00 | 306 | 15 | 333.74 | < .0001 | < .0001 |
|  | +09 -48 +30 | 98 | 8 | 330.19 | < .0001 | < .0001 |
|  | +63 -09 +30 | 480 | 41 | 316.08 | < .0001 | < .0001 |
|  | +66 -12 +00 | 4 | 1 | 167.70 | .0380 | .0030 |
|  | -63 -03 +24 | 9 | 1 | 163.00 | .0470 | .0035 |
|  | +27 +27 -03 | 1 | 1 | 160.93 | .0500 | .0038 |
| **Supplementary Table S3.** Voxel-wise comparisons between ASD and TD regarding LCOR in the ABIDE2 cohort. List of significant TFCE-corrected cluster for positive (ASD>TD) and negative contrast (ASD<TD); highlighted cluster coordinates indicate the voxel with the highest T-value within a cluster; ABIDE = autism brain imaging data exchange; ASD = autism spectrum disorder; TD = typically developed controls; TFCE = threshold free cluster enhancement; FDR = false discovery rate; FWE = family-wise error; LCOR = local synchronization; TFCE-threshold: *p* < 0.05. | | | | | | |

Contrast: ASD>TD

42 voxels (31%) covering 3% of atlas.Brain-Stem

14 voxels (10%) covering 2% of atlas.TP r (Temporal Pole Right)

13 voxels (9%) covering 7% of atlas.SMA r (Supplementary Motor Cortex- Right)

12 voxels (9%) covering 2% of atlas.Cereb8 r (Cerebelum 8 Right)

11 voxels (8%) covering 2% of atlas.TP l (Temporal Pole Left)

6 voxels (4%) covering 6% of atlas.aITG r (Inferior Temporal Gyrus, anterior division Right)

5 voxels (4%) covering 2% of atlas.Cereb9 l (Cerebelum 9 Left)

3 voxels (2%) covering 1% of atlas.pSMG r (Supramarginal Gyrus, posterior division Right)

3 voxels (2%) covering 1% of atlas.Cereb8 l (Cerebelum 8 Left)

2 voxels (1%) covering 2% of atlas.aTFusC r (Temporal Fusiform Cortex, anterior division Right)

2 voxels (1%) covering 1% of atlas.pTFusC r (Temporal Fusiform Cortex, posterior division Right)

1 voxels (1%) covering 0% of atlas.toMTG l (Middle Temporal Gyrus, temporooccipital part Left)

1 voxels (1%) covering 0% of atlas.pITG r (Inferior Temporal Gyrus, posterior division Right)

1 voxels (1%) covering 0% of atlas.AG r (Angular Gyrus Right)

21 voxels (15%) covering 0% of atlas.not-labeled

Contrast: ASD<TD

105 voxels (12%) covering 26% of atlas.IC r (Insular Cortex Right)

100 voxels (11%) covering 10% of atlas.PostCG r (Postcentral Gyrus Right)

83 voxels (9%) covering 12% of atlas.PC (Cingulate Gyrus, posterior division)

76 voxels (8%) covering 6% of atlas.PreCG r (Precentral Gyrus Right)

72 voxels (8%) covering 19% of atlas.PaCiG r (Paracingulate Gyrus Right)

62 voxels (7%) covering 3% of atlas.FP r (Frontal Pole Right)

62 voxels (7%) covering 17% of atlas.PaCiG l (Paracingulate Gyrus Left)

56 voxels (6%) covering 21% of atlas.CO r (Central Opercular Cortex Right)

47 voxels (5%) covering 6% of atlas.AC (Cingulate Gyrus, anterior division)

28 voxels (3%) covering 12% of atlas.aSMG r (Supramarginal Gyrus, anterior division Right)

26 voxels (3%) covering 1% of atlas.FP l (Frontal Pole Left)

25 voxels (3%) covering 16% of atlas.PO r (Parietal Operculum Cortex Right)

17 voxels (2%) covering 6% of atlas.MedFC (Frontal Medial Cortex)

15 voxels (2%) covering 3% of atlas.FOrb r (Frontal Orbital Cortex Right)

13 voxels (1%) covering 1% of atlas.Precuneous (Precuneous Cortex)

12 voxels (1%) covering 9% of atlas.PT r (Planum Temporale Right)

11 voxels (1%) covering 12% of atlas.FO r (Frontal Operculum Cortex Right)

9 voxels (1%) covering 4% of atlas.IFG oper r (Inferior Frontal Gyrus, pars opercularis Right)

4 voxels (0%) covering 0% of atlas.PreCG l (Precentral Gyrus Left)

4 voxels (0%) covering 0% of atlas.PostCG l (Postcentral Gyrus Left)

3 voxels (0%) covering 0% of atlas.TP r (Temporal Pole Right)

2 voxels (0%) covering 2% of atlas.pSTG r (Superior Temporal Gyrus, posterior division Right)

2 voxels (0%) covering 2% of atlas.HG r (Heschl's Gyrus Right)

2 voxels (0%) covering 1% of atlas.Putamen r

62 voxels (7%) covering 0% of atlas.not-labeled

**Supplementary Table S4.** Regions with significant voxels in ABIDE2 (all clusters combined). List of all significant voxels and atlas regions in the autism brain imaging data exchange (ABIDE) 1 dataset for the positive (ASD>TD) and negative contrast (ASD<TD); ASD = autism spectrum disorder; TD = typically developed controls.

**Supplementary Table S5. LCOR alterations in ASD relate to the *in-vivo* distribution of neurotransmitter systems**

|  | **Co-localization** | **ABIDE1** | |  | **ABIDE2** | |
| --- | --- | --- | --- | --- | --- | --- |
|  |  | **Spearman *r*** | ***p*-value** |  | **Spearman *r*** | ***p*-value** |
| **LCOR** | 5HT1a | .1982 | .1349 |  | .0492 | .7213 |
|  | 5HT1b | -.2480 | .0809 |  | -.4308 | < .0010*** |
|  | 5HT2a | .0555 | .6993 |  | -.0002 | .9980 |
|  | 5HT4 | .1945 | .0240* |  | -.0334 | .7423 |
|  | SERT | -.0393 | .6553 |  | -.0245 | .7982 |
|  | D1 | -.2634 | .0050** |  | -.2778 | .0020** |
|  | D2 | -.3216 | < .0010*** |  | -.3239 | < .0010*** |
|  | DAT | -.1867 | .0330* |  | -.1963 | .0280* |
|  | FDOPA | -.2000 | .0340* |  | -.0794 | .3996 |
|  | NMDA | -.3318 | .0090** |  | -.3109 | .0090** |
|  | mGluR5 | -.3786 | .0040** |  | -.3923 | .0030** |
|  | GABAa | -.2478 | .0050** |  | -.1864 | .0350* |
|  | CB1 | -.1307 | .3816 |  | -.3348 | .0110* |
|  | MU | -.1056 | .4905 |  | -.1484 | .3400 |
|  | NAT | -.2018 | .0240* |  | -.1641 | .0639 |
|  | VAChT | -.2409 | .0030** |  | -.2378 | .0060** |
|  | CBF | -.3507 | .0130** |  | -.3595 | .0060** |
| **Supplementary Table S5.** Statistical data of co-localizations between LCOR alterations in ASD compared to TD and neurotransmitter systems in ABIDE1 and ABIDE2. ABIDE = autism brain imaging data exchange; ASD = autism spectrum disorder; LCOR = local synchronization; TD = typically developed controls; 5HT = 5-hydroxytryptamine (serotonin receptor); SERT = serotonin transporter; D1/D2 = dopamine receptor; DAT = dopamine transporter; FDOPA = fluorodopa; NMDA = N-Methyl-D-Aspartat receptor; mGluR5 = metabotropic glutamate receptor; GABAa = γ-aminobutyric acid type A receptor; CB1 = cannabinoid receptor; MU = μ-opioid receptor; NAT = noradrenaline transporter; VAChT = vesicular acetylcholine transporter; CBF = cerebral blood flow; * = *p* < .05; ** = *p* < .01; *** = *p* < .001. | | | | | | |

In ABIDE1, we found significant co-localizations between LCOR alterations in ASD compared to TD and serotonergic 5HT4, dopaminergic D1, D2, DAT and FDOPA, glutamatergic NMDA and mGLuR5, GABAa, NAT and VAChT distributions from nuclear imaging. The significant co-localizations with D1, D2, DAT, NMDA, GluR5, NMDA and VAChT were replicated in the ABIDE2 dataset.

**Supplementary Table S6. Neurotransmitter co-localizations with LCOR changes induced by ketamine and midazolam**

|  | **Co-localization** | **Ketamine** | |  | **Midazolam** | |
| --- | --- | --- | --- | --- | --- | --- |
|  |  | **Spearman *r*** | ***p-FDR*** |  | **Spearman *r*** | ***p-FDR*** |
| **LCOR** | 5HT1a | .0487 | .8460 |  | -.1613 | .2652 |
|  | 5HT1b | -.3265 | .1199 |  | -.2478 | .1191 |
|  | 5HT2a | -.2299 | .2420 |  | -.3052 | .0466* |
|  | 5HT4 | -.0487 | .7353 |  | -.0824 | .4018 |
|  | SERT | -.1261 | .3149 |  | .4363 | < .001*** |
|  | D1 | -.2829 | .0320* |  | .0960 | .3418 |
|  | D2 | -.0265 | .8109 |  | .1757 | .1052 |
|  | DAT | -.0270 | .7752 |  | .4259 | .0016** |
|  | FDOPA | -.1336 | .3536 |  | .4169 | < .001*** |
|  | NMDA | -.3380 | .0320* |  | .2254 | .1187 |
|  | mGluR5 | -.2926 | .1375 |  | -.1691 | .2797 |
|  | GABAa | -.2514 | .0373* |  | -.1760 | .0962 |
|  | CB1 | -.2657 | .2504 |  | -.4683 | .0022** |
|  | MU | -.1732 | .5740 |  | -.1212 | .5067 |
|  | NAT | .0778 | .5395 |  | .2639 | .0096** |
|  | VAChT | -.1236 | .3481 |  | .3324 | < .001*** |
| **Supplementary Table S6.** Statistical data of co-localizations between LCOR alterations induced by ketamine and midazolam compared to placebo and neurotransmitter systems LCOR = local synchronization; FDR = false discovery rate; 5HT = 5-hydroxytryptamine (serotonin receptor); SERT = serotonin transporter; D1/D2 = dopamine receptor; DAT = dopamine transporter; FDOPA = fluorodopa; NMDA = N-Methyl-D-Aspartat receptor; mGluR5 = metabotropic glutamate receptor; GABAa = γ-aminobutyric acid type A receptor; CB1 = cannabinoid receptor; MU = μ-opioid receptor; NAT = noradrenaline transporter; VAChT = vesicular acetylcholine transporter; CBF = cerebral blood flow; * = *p* < .05; ** = *p* < .01; *** = *p* < .001. | | | | | | |

We computed spatial Spearman correlations between the whole-brain LCOR T-maps induced by ketamine or midazolam and different neurotransmitter distributions. The ketamine effect on LCOR was co-localized with the distribution of D1, NMDA and GABAa receptors after correcting for multiple comparisons. Midazolam displayed significant co-localizations with 5HT2a, SERT, DAT, FDOPA, CB1, NAT and VAChT.

**Supplementary Fig. S1. Visualization of ketamine and midazolam co-localization profiles**

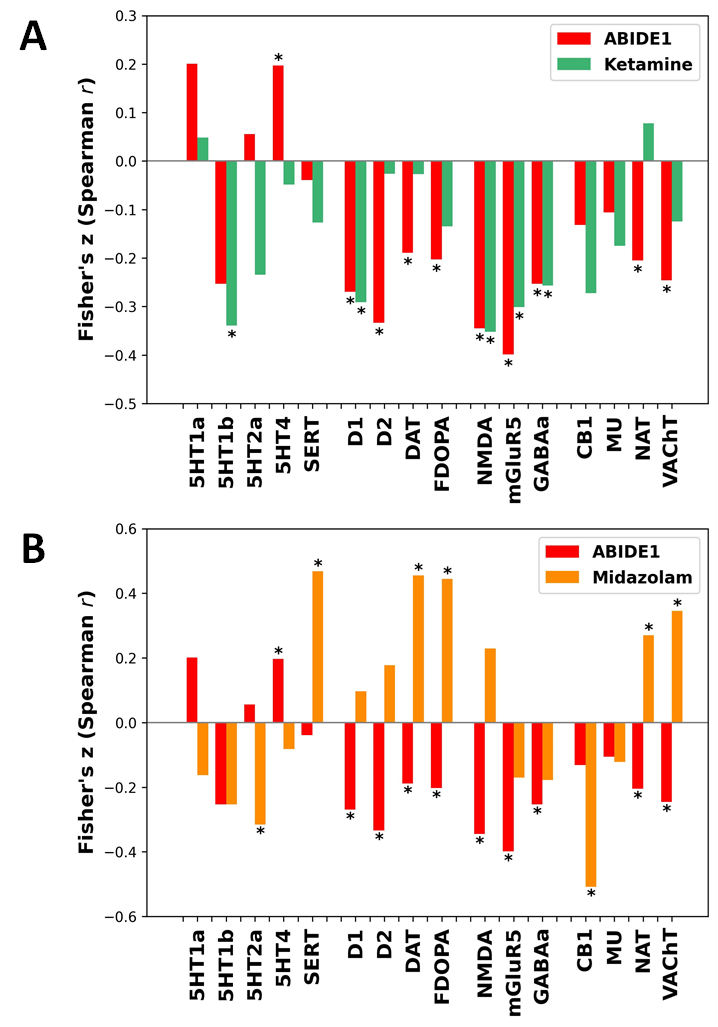

**Supplementary Fig. S1.** Neurotransmitter co-localizations with LCOR changes induced by ketamine and midazolam compared to the co-localization profile of ASD in ABIDE1. (A) Neurotransmitter co-localizations with local synchronization (LCOR) alterations induced by NMDA antagonist ketamine (versus placebo) compared to the co-localization profile in autism spectrum disorder (versus typically developed controls) in ABIDE1. (B) Neurotransmitter co-localizations with LCOR alterations induced by GABAa potentiator midazolam (versus placebo) compared to the co-localization profile in autism spectrum disorder (versus typically developed controls) in ABIDE1. The asterisk (*) represents significant co-localizations (*p* < .05).

**Supplementary Table S7-10. LCOR alterations induced by ketamine and midazolam administration**

We computed voxel-wise whole-brain comparisons of LCOR induced by ketamine and midazolam administration compared to placebo within the CONN toolbox^60^.

| **contrast** | **cluster (x, y, z)** | **size** | **peaks** | **TFCE** | **peak *p-FWE*** | **peak *p-FDR*** |
| --- | --- | --- | --- | --- | --- | --- |
| **KET>PLC** | +22 -14 -36 | 847 | 20 | 925.07 | .0010 | .0012 |
|  | +24 -42 -48 | 783 | 17 | 875.15 | .0030 | .0012 |
|  | +38 -52 -30 | 90 | 4 | 730.68 | .0110 | .0012 |
|  | -34 -82 +12 | 106 | 1 | 723.52 | .0110 | .0012 |
|  | -40 -52 -42 | 97 | 1 | 719.73 | .0120 | .0012 |
|  | +28 -46 -16 | 95 | 4 | 712.09 | .0120 | .0012 |
|  | +02 -82 -42 | 107 | 2 | 661.03 | .0230 | .0017 |
|  | +52 +00 +14 | 28 | 1 | 640.31 | .0320 | .0021 |
|  | -10 -72 -48 | 20 | 1 | 625.02 | .0350 | .0021 |
|  | -24 -64 -50 | 46 | 3 | 619.83 | .0390 | .0021 |
| **KET<PLC** | -06 -28 +62 | 2302 | 29 | -1596.81 | < .0001 | < .0001 |
|  | -36 +40 +02 | 449 | 11 | -989.32 | .0020 | .0014 |
|  | +50 +26 -14 | 368 | 9 | -941.97 | .0020 | .0014 |
|  | +08 +32 +22 | 339 | 9 | -904.17 | .0030 | .0015 |
|  | -50 +10 -08 | 99 | 4 | -631.31 | .0380 | .0025 |
|  | -34 +12 -04 | 67 | 2 | -630.59 | .0390 | .0025 |
|  | +44 +44 -18 | 13 | 1 | -611.86 | .0430 | .0026 |
|  | +34 +58 -18 | 29 | 1 | -595.40 | .0470 | .0028 |
| **Supplementary Table S7.** Main effect of ketamine on LCOR compared to placebo condition. List of significant TFCE-corrected cluster for positive (ketamine>placebo) and negative contrast (ketamine<placebo); highlighted cluster coordinates indicate the voxel with the highest T-value within a cluster; KET = ketamine; PLC = placebo; TFCE = threshold free cluster enhancement; FDR = false discovery rate; FWE = family-wise error; LCOR = local synchronization; TFCE-threshold: *p* < 0.05. | | | | | | |

Ketamine induced LCOR reductions in pre- and postcentral gyri, frontal cortices, precuneus, cingulate, insular and opercular cortices, whereas LCOR was increased particularly in the cerebellum and temporal regions. Midazolam induced LCOR reductions in the left lateral occipital cortex and increases within the left hemispheric supplementary motor cortex, precentral gyrus, putamen, insular cortex and middle temporal gyrus as well as anterior cingulate cortex.

Contrast: KET>PLC

286 voxels (13%) covering 36% of atlas.Cereb9 r (Cerebelum 9 Right)

167 voxels (8%) covering 57% of atlas.aTFusC r (Temporal Fusiform Cortex, anterior division Right)

163 voxels (7%) covering 7% of atlas.Cereb8 r (Cerebelum 8 Right)

128 voxels (6%) covering 5% of atlas.TP r (Temporal Pole Right)

111 voxels (5%) covering 17% of atlas.aPaHC r (Parahippocampal Gyrus, anterior division Right)

102 voxels (5%) covering 12% of atlas.Cereb9 l (Cerebelum 9 Left)

94 voxels (4%) covering 4% of atlas.Cereb1 l (Cerebelum Crus1 Left)

92 voxels (4%) covering 2% of atlas.sLOC l (Lateral Occipital Cortex, superior division Left)

77 voxels (3%) covering 9% of atlas.TOFusC r (Temporal Occipital Fusiform Cortex Right)

68 voxels (3%) covering 2% of atlas.Brain-Stem

67 voxels (3%) covering 4% of atlas.Cereb8 l (Cerebelum 8 Left)

51 voxels (2%) covering 16% of atlas.aITG r (Inferior Temporal Gyrus, anterior division Right)

48 voxels (2%) covering 2% of atlas.Cereb2 r (Cerebelum Crus2 Right)

44 voxels (2%) covering 3% of atlas.Cereb6 r (Cerebelum 6 Right)

39 voxels (2%) covering 5% of atlas.pTFusC r (Temporal Fusiform Cortex, posterior division Right)

28 voxels (1%) covering 1% of atlas.Cereb1 r (Cerebelum Crus1 Right)

27 voxels (1%) covering 17% of atlas.Cereb10 r (Cerebelum 10 Right)

16 voxels (1%) covering 1% of atlas.LG r (Lingual Gyrus Right)

12 voxels (1%) covering 2% of atlas.Cereb7 r (Cerebelum 7b Right)

9 voxels (0%) covering 0% of atlas.PreCG r (Precentral Gyrus Right)

7 voxels (0%) covering 1% of atlas.Cereb7 l (Cerebelum 7b Left)

6 voxels (0%) covering 2% of atlas.Amygdala r

3 voxels (0%) covering 0% of atlas.Hippocampus r

2 voxels (0%) covering 0% of atlas.Cereb2 l (Cerebelum Crus2 Left)

572 voxels (26%) covering 0% of atlas.not-labeled

Contrast: KET<PLC

839 voxels (23%) covering 20% of atlas.PreCG r (Precentral Gyrus Right)

491 voxels (13%) covering 11% of atlas.PreCG l (Precentral Gyrus Left)

396 voxels (11%) covering 6% of atlas.FP l (Frontal Pole Left)

331 voxels (9%) covering 10% of atlas.PostCG r (Postcentral Gyrus Right)

308 voxels (8%) covering 12% of atlas.AC (Cingulate Gyrus, anterior division)

183 voxels (5%) covering 5% of atlas.PostCG l (Postcentral Gyrus Left)

150 voxels (4%) covering 10% of atlas.FOrb r (Frontal Orbital Cortex Right)

88 voxels (2%) covering 7% of atlas.IC l (Insular Cortex Left)

83 voxels (2%) covering 1% of atlas.FP r (Frontal Pole Right)

50 voxels (1%) covering 2% of atlas.TP l (Temporal Pole Left)

33 voxels (1%) covering 1% of atlas.TP r (Temporal Pole Right)

25 voxels (1%) covering 2% of atlas.PaCiG r (Paracingulate Gyrus Right)

18 voxels (0%) covering 0% of atlas.Precuneous (Precuneous Cortex)

16 voxels (0%) covering 3% of atlas.IFG tri r (Inferior Frontal Gyrus, pars triangularis Right)

16 voxels (0%) covering 2% of atlas.IFG oper r (Inferior Frontal Gyrus, pars opercularis Right)

12 voxels (0%) covering 1% of atlas.PC (Cingulate Gyrus, posterior division)

4 voxels (0%) covering 0% of atlas.CO l (Central Opercular Cortex Left)

3 voxels (0%) covering 0% of atlas.MidFG l (Middle Frontal Gyrus Left)

3 voxels (0%) covering 1% of atlas.FO l (Frontal Operculum Cortex Left)

2 voxels (0%) covering 0% of atlas.IC r (Insular Cortex Right)

2 voxels (0%) covering 0% of atlas.SMA r (Juxtapositional Lobule Cortex -formerly Supplementary Motor Cortex- Right)

1 voxels (0%) covering 0% of atlas.IFG tri l (Inferior Frontal Gyrus, pars triangularis Left)

1 voxels (0%) covering 0% of atlas.PaCiG l (Paracingulate Gyrus Left)

1 voxels (0%) covering 0% of atlas.FO r (Frontal Operculum Cortex Right)

610 voxels (17%) covering 0% of atlas.not-labeled

**Supplementary Table S8.** Regions with significant LCOR alterations induced by ketamine (all clusters combined). List of all significant voxels and atlas regions with altered LCOR induced by ketamine compared to placebo for the positive (KET>PLC) and negative contrast (KET<PLC); KET = ketamine; PLC = placebo; LCOR = local synchronization.

| **contrast** | **cluster (x, y, z)** | **size** | **peaks** | **TFCE** | **peak p-FWE** | **peak p-FDR** |
| --- | --- | --- | --- | --- | --- | --- |
| **MDZ>PLC** | -08 -14 +44 | 83 | 3 | 685.91 | .0330 | .0184 |
|  | -32 -02 +02 | 20 | 1 | 678.34 | .0330 | .0184 |
|  | -46 -20 -14 | 37 | 1 | 657.11 | .0430 | .0184 |
| **MDZ<PLC** | -34 -74 +34 | 120 | 1 | -821.65 | .0060 | .0188 |
| **Supplementary Table S9.** Main effect of midazolam on LCOR compared to placebo condition. List of significant TFCE-corrected cluster for positive (midazolam>placebo) and negative contrast (midazolam<placebo); highlighted cluster coordinates indicate the voxel with the highest T-value within a cluster; MDZ = midazolam; PLC = placebo; TFCE = threshold free cluster enhancement; FDR = false discovery rate; FWE = family-wise error; LCOR = local synchronization; TFCE-threshold: *p* < 0.05. | | | | | | |

Contrast: MDZ>PLC

29 voxels (21%) covering 5% of atlas.SMA L (Supplementary Motor Cortex- Left)

20 voxels (14%) covering 0% of atlas.PreCG l (Precentral Gyrus Left)

7 voxels (5%) covering 1% of atlas.Putamen l

4 voxels (3%) covering 0% of atlas.IC l (Insular Cortex Left)

4 voxels (3%) covering 0% of atlas.pMTG l (Middle Temporal Gyrus, posterior division Left)

3 voxels (2%) covering 0% of atlas.AC (Cingulate Gyrus, anterior division)

1 voxels (1%) covering 0% of atlas.PP l (Planum Polare Left)

72 voxels (51%) covering 0% of atlas.not-labeled

Contrast: MDZ<PLC

120 voxels (100%) covering 2% of atlas.sLOC l (Lateral Occipital Cortex, superior division Left)

**Supplementary Table S10.** Regions with significant LCOR alterations induced by midazolam (all clusters combined). List of all significant voxels and atlas regions with altered LCOR induced by midazolam compared to placebo for the positive (MDZ>PLC) and negative contrast (MDZ<PLC); MDZ = midazolam; PLC = placebo; LCOR = local synchronization.

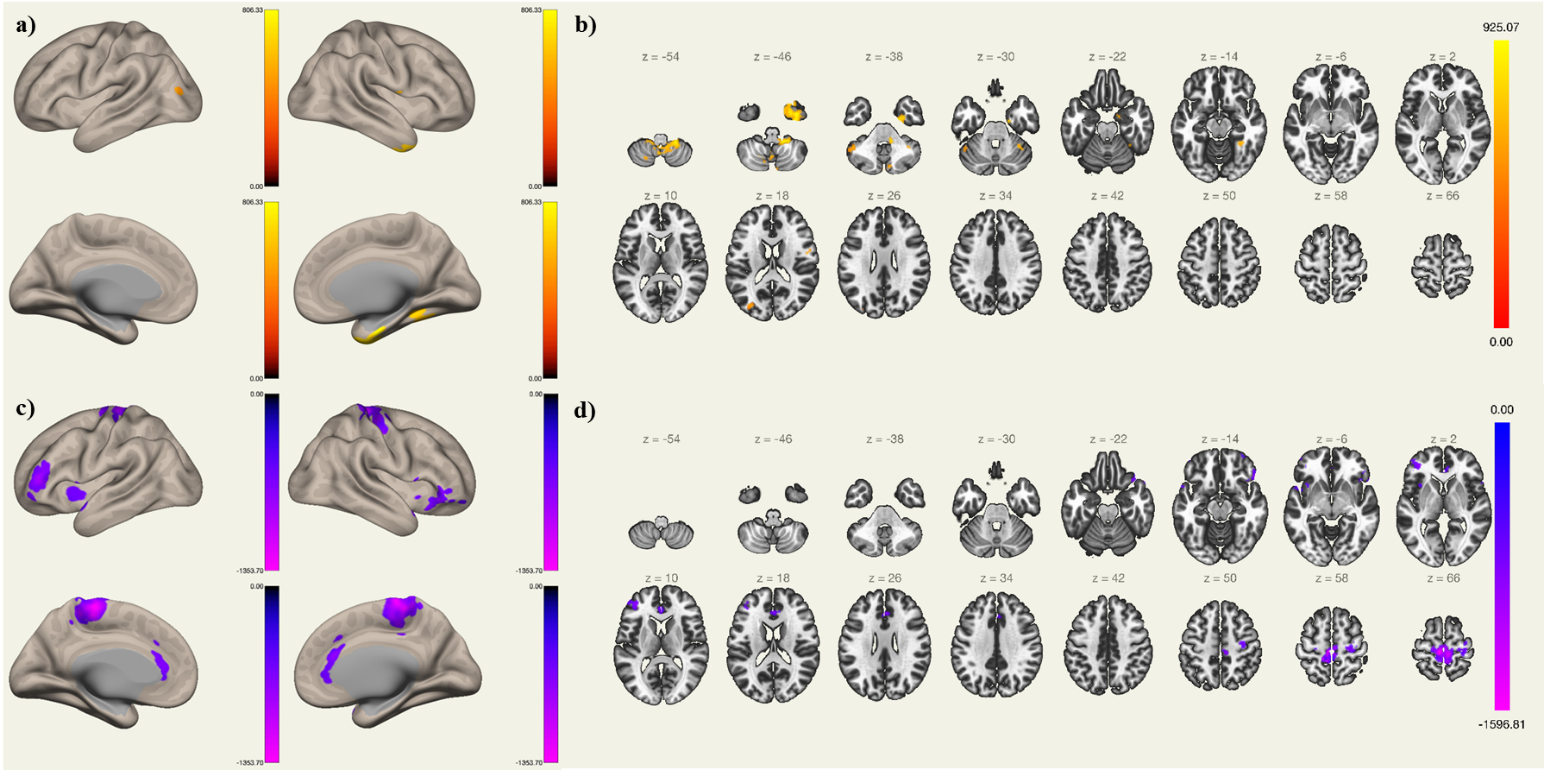
**Supplementary Fig. S2-3. Visualization of ketamine and midazolam induced LCOR changes**

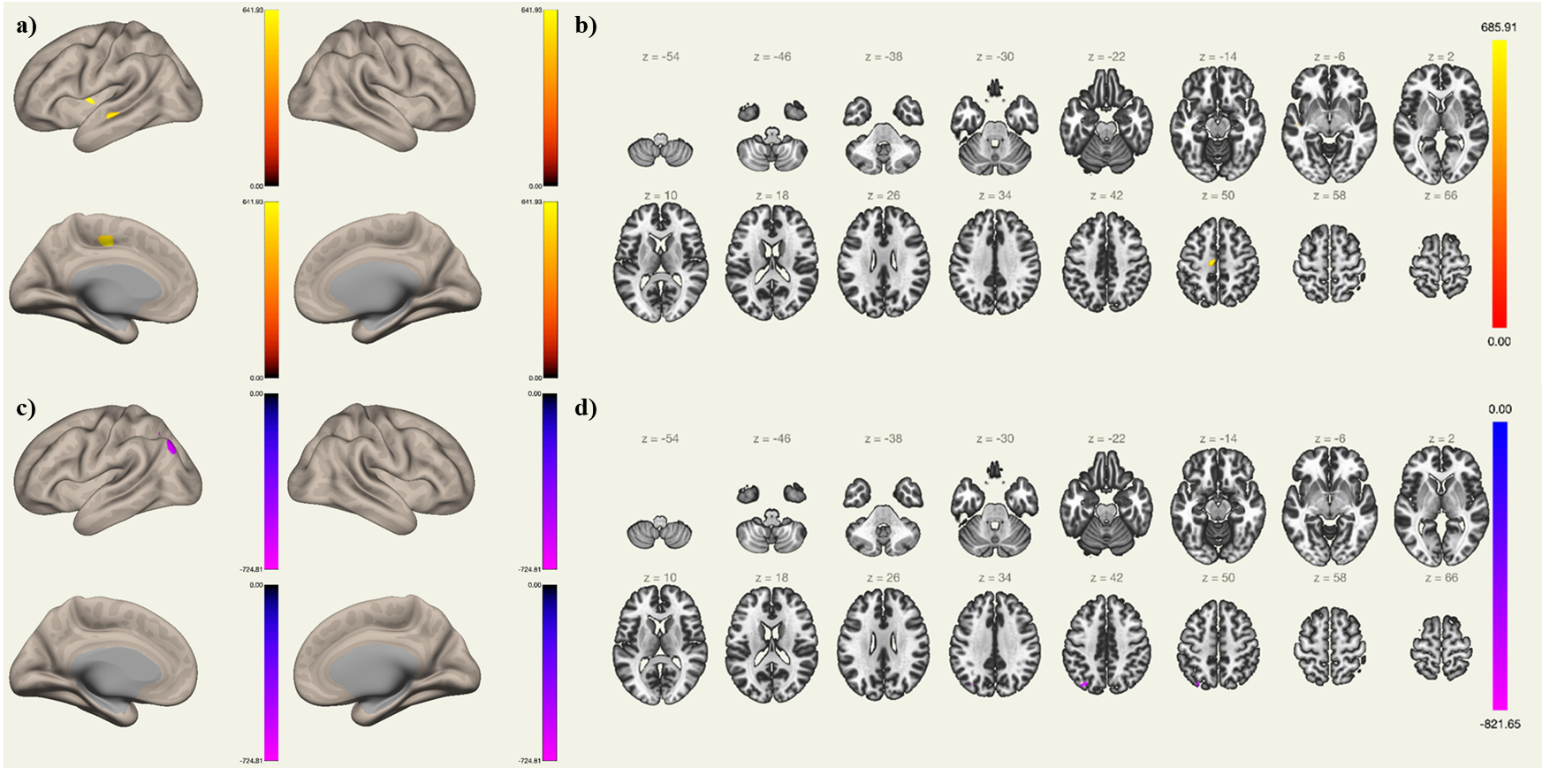
**Supplementary Fig. S2.** Significant voxel-wise LCOR changes after ketamine administration compared to placebo condition. Increased LCOR induced by ketamine compared to placebo (red-yellow), a) sagittal view and b) axial view. Decreased LCOR after ketamine administration compared to placebo (blue-purple), c) sagittal view and d) axial view.

**Supplementary Fig. S3.** Significant voxel-wise LCOR changes after midazolam administration compared to placebo condition. Increased LCOR induced by midazolam compared to placebo (red-yellow), a) sagittal view and b) axial view. Decreased LCOR after midazolam administration compared to placebo (blue-purple), c) sagittal view and d) axial view.

**Supplementary Table S11. Similarity of ASD and medication effects**

To assess the similarity between the brain pattern observed in ASD and the LCOR alterations induced by ketamine and midazolam, we conducted a correlation analysis within the JuSpace toolbox^65^ using LCOR T-maps generated by the CONN toolbox^60^. Our findings reveal a significant positive correlation between ketamine-induced and ASD-related global brain patterns, indicating that ketamine elicits ASD-like local activity patterns. In contrast, midazolam did not show a similar correlation.

Furthermore, while the whole-brain LCOR T-maps for ketamine and midazolam were not significantly correlated with each other, the LCOR patterns observed in the ABIDE1 and ABIDE2 datasets demonstrated a strong correlation.

|  | **Pearson *r*** | ***p*** |
| --- | --- | --- |
| ABIDE I vs. KET | .381 | .003** |
| ABIDE I vs. MDZ | .018 | .903 |
| ABIDE II vs. KET | .292 | .036* |
| ABIDE II vs. MDZ | .205 | .155 |
| ABIDE I vs. ABIDE II | .597 | < .001*** |
| KET vs. MDZ | .075 | .637 |
| **Supplementary Table S11.** Correlation between ASD-specific and pharmaco-induced T-maps. ABIDE = Autism Brain Imaging Data Exchange; ASD = autism spectrum disorder; KET = ketamine; MDZ = midazolam; ASD contrast (ASD>TD) vs. ketamine contrast (KET>PLC) vs. midazolam contrast (MDZ>TD). | | |

**Supplementary** **Table S12. Correlation of the LCOR-neurotransmitter co-localizations with ASD symptom domains in ABIDE1 and ABIDE2**

We extracted individual Fisher’s z scores for all subjects with ASD from ABIDE1 and ABIDE2 separately and computed Pearson correlation analyses with the autism diagnostic observation schedule (ADOS) subscales^67^. For the consistent significant LCOR-neurotransmitter co-localizations observed across both ABIDE datasets, we did not find any significant association with clinical symptom domains in ABIDE1. The co-localization with GABAa and VAChT was significantly associated with stereotyped behavior and restricted interests (SBRI) in ABIDE2.

|  |  | **Communication** | |  | **Social interaction** | |  | **SBRI** | |
| --- | --- | --- | --- | --- | --- | --- | --- | --- | --- |
|  |  | *r* | *p* |  | *r* | *p* |  | *r* | *p* |
| 5HT1a | ABIDE1 | -.013 | .821 |  | -.014 | .806 |  | -.016 | .806 |
|  | ABIDE2 | .009 | .889 |  | -.026 | .692 |  | .016 | .809 |
| 5HT1b | ABIDE1 | .172 | .029* |  | -.049 | .394 |  | .009 | .893 |
|  | ABIDE2 | -.055 | .401 |  | .036 | .578 |  | -.046 | .483 |
| 5HT2a | ABIDE1 | .047 | .421 |  | -.052 | .363 |  | .026 | .685 |
|  | ABIDE2 | -.062 | .342 |  | -.047 | .470 |  | -.088 | .176 |
| 5HT4 | ABIDE1 | -.057 | .324 |  | -.045 | .440 |  | -.025 | .701 |
|  | ABIDE2 | -.147 | .024* |  | -.053 | .411 |  | .044 | .495 |
| SERT | ABIDE1 | -.116 | .046* |  | .003 | .964 |  | -.109 | .087 |
|  | ABIDE2 | -.110 | .089 |  | -.038 | .559 |  | .034 | .601 |
| D1 | ABIDE1 | -.047 | .417 |  | .010 | .857 |  | -.016 | .797 |
|  | ABIDE2 | -.095 | .146 |  | .046 | .481 |  | .113 | .083 |
| D2 | ABIDE1 | -.016 | .777 |  | -.009 | .878 |  | -.046 | .476 |
|  | ABIDE2 | -.078 | .233 |  | .007 | .909 |  | -.015 | .820 |
| DAT | ABIDE1 | -.055 | .346 |  | .046 | .427 |  | -.061 | .341 |
|  | ABIDE2 | -.071 | .273 |  | .019 | .768 |  | .105 | .105 |
| FDOPA | ABIDE1 | -.060 | .303 |  | .061 | .286 |  | -.071 | .264 |
|  | ABIDE2 | -.042 | .522 |  | .024 | .712 |  | .065 | .317 |
| NMDA | ABIDE1 | -.032 | .580 |  | .011 | .849 |  | -.112 | .080 |
|  | ABIDE2 | -.092 | .155 |  | -.023 | .730 |  | -.039 | .548 |
| mGluR5 | ABIDE1 | .035 | .552 |  | -.027 | .644 |  | .021 | .746 |
|  | ABIDE2 | -.022 | .733 |  | .050 | .445 |  | -.065 | .321 |
| GABAa | ABIDE1 | .077 | .185 |  | -.044 | .445 |  | .028 | .663 |
|  | ABIDE2 | -.014 | .833 |  | -.004 | .948 |  | -.133 | .040* |
| CB1 | ABIDE1 | .049 | .401 |  | -.043 | .456 |  | .073 | .253 |
|  | ABIDE2 | -.031 | .630 |  | .063 | .330 |  | < .001 | .996 |
| MU | ABIDE1 | < .001 | .998 |  | -.005 | .933 |  | .033 | .608 |
|  | ABIDE2 | -.014 | .836 |  | .097 | .136 |  | .171 | .008** |
| NAT | ABIDE1 | .075 | .199 |  | .094 | .101 |  | -.057 | .369 |
|  | ABIDE2 | .028 | .668 |  | -.049 | .454 |  | -.044 | .502 |
| VAChT | ABIDE1 | -.034 | .555 |  | .076 | .189 |  | -.032 | .613 |
|  | ABIDE2 | -.027 | .679 |  | .040 | .536 |  | .141 | .030* |
| **Supplementary Table S12.** Pearson correlations between the strength of LCOR-neurotransmitter co-localizations in ASD and the ADOS subscales. ADOS = autism diagnostic observation schedule; ASD = autism spectrum disorder; LCOR = local synchronization; SBRI = stereotyped behavior and restricted interests; 5HT = 5-hydroxytryptamine (serotonin receptor); SERT = serotonin transporter; D1/D2 = dopamine receptor; DAT = dopamine transporter; FDOPA = fluorodopa; NMDA = N-Methyl-D-Aspartat receptor; mGluR5 = metabotropic glutamate receptor; GABAa = γ-aminobutyric acid type A receptor; CB1 = cannabinoid receptor; MU = μ-opioid receptor; NAT = noradrenaline transporter; VAChT = vesicular acetylcholine transporter; CBF = cerebral blood flow; * = *p* < .05; ** = *p* < .01; *** = *p* < .001. | | | | | | | | | |

**Supplementary** **Fig. S4. Correlation of the LCOR-neurotransmitter co-localizations in ASD with clinical symptom domains**

There was no significant association between the consistent LCOR-neurotransmitter co-localizations and clinical symptom severity as measures with the ADOS score in the ABIDE1 dataset. A significant but weak negative association was observed in ABIDE2 between SBRI and the strength of LCOR-GABAa co-localizations (*r* = -.133, *p* = .040) and a positive with the strength of LCOR-VAChT co-localizations (*r* = .141, *p* = .030). Both findings did not survive a correction for multiple comparisons.

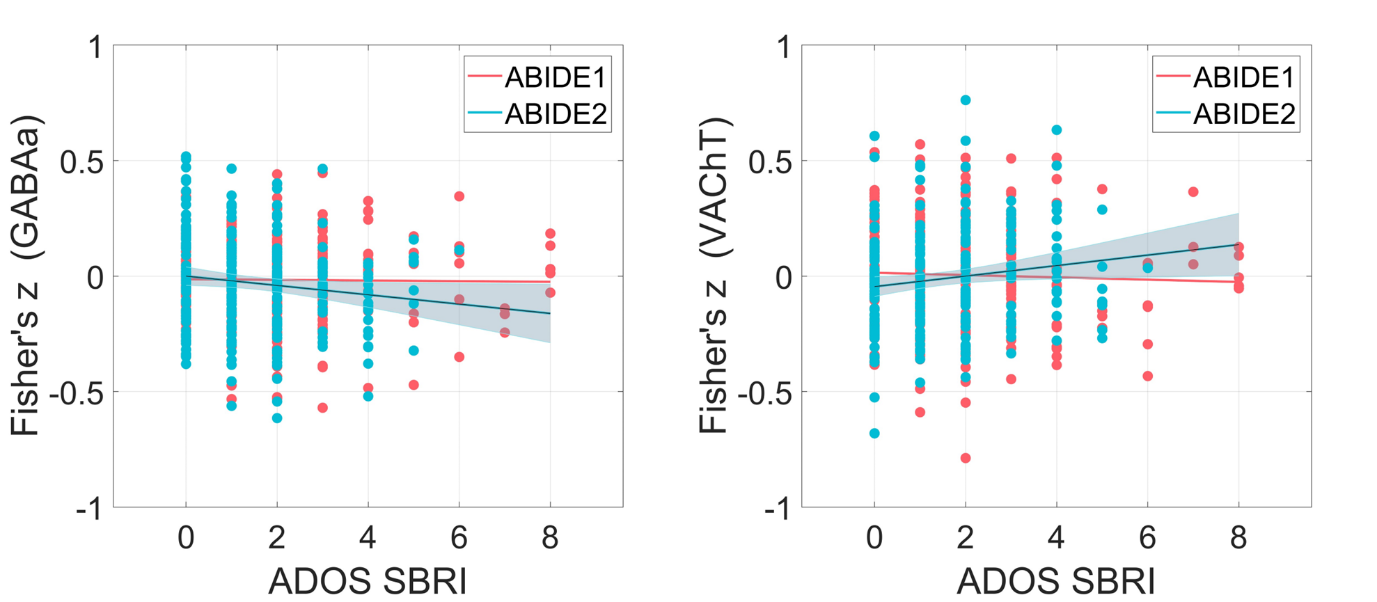

**Supplementary Fig. S4.** Association of the co-localizations between LCOR and GABAa and VAChT distributions with stereotyped behavior and restricted interests (SBRI). Scatterplot showing values of the SBRI subscale of the Autism Diagnostic Observation Schedule (ADOS) relative to the strength of the spatial co-localization (individual Fisher’s z-scores) between local synchronization (LCOR) and GABAa as well as VAChT distributions in subjects with autism spectrum disorder (ASD).

**Materials and Methods**

**Supplementary Fig. S5. Exclusion process**

We excluded subjects with intellectual disability (IQ ≤ 70), subjects with missing information and subjects with excessive head motion during image acquisition (translation > 3 mm or rotation > 3°). For ABIDE2, another 3 subjects were excluded due to preprocessing failure of functional imaging data.

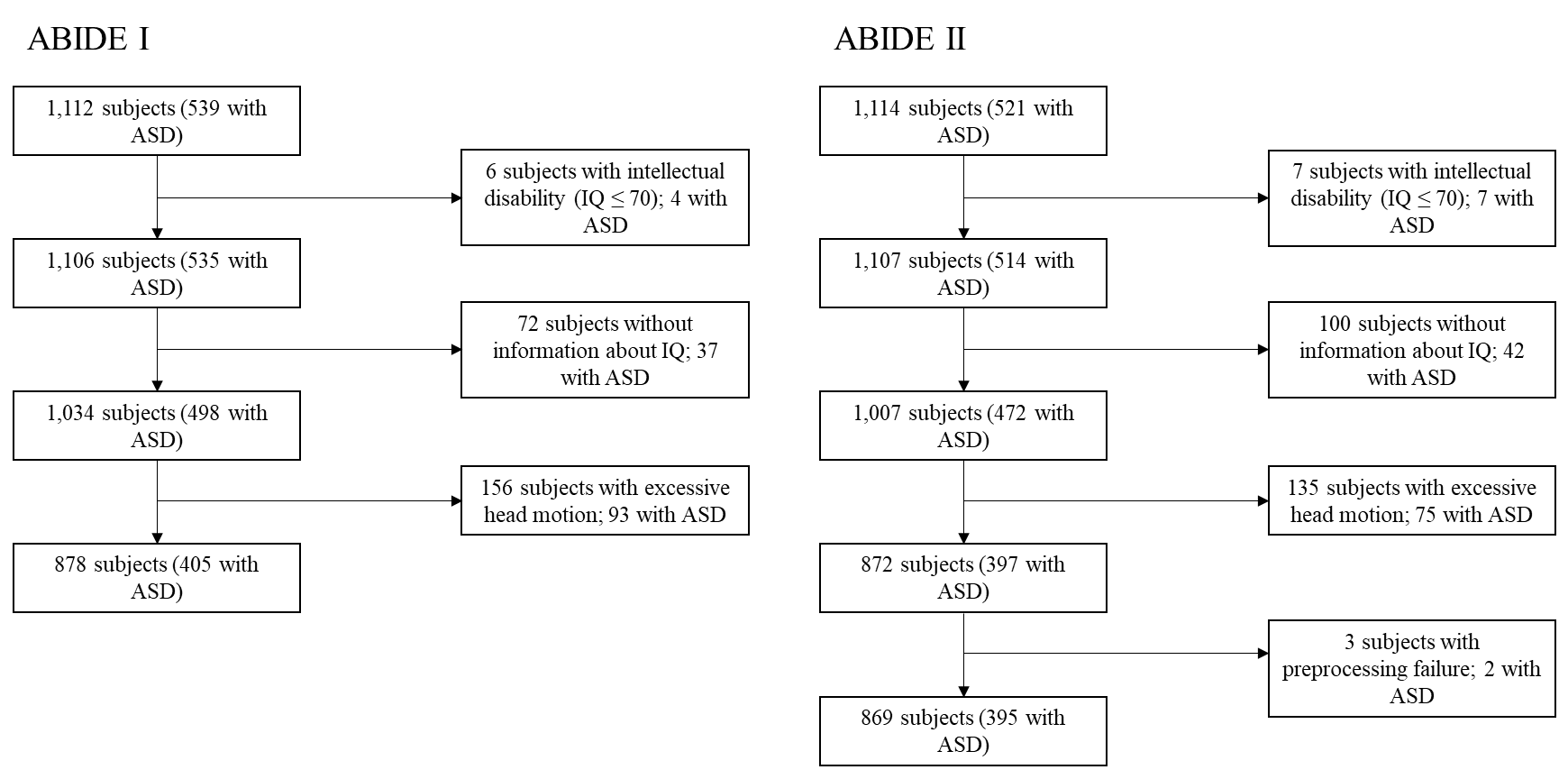

**Supplementary Fig. S5.** Overview of the exclusion process for the Autism Brain Imaging Data Exchange (ABIDE) 1 and ABIDE2 datasets, ASD = autism spectrum disorder, IQ = intelligence quotient.

**Supplementary Table S13-14. MRI data acquisition**

| **Supplementary Table S13.** Detailed description of MRI acquisition parameters of the respective sites included in ABIDE I. | | | | | | | | | | | | |
| --- | --- | --- | --- | --- | --- | --- | --- | --- | --- | --- | --- | --- |
| **Scanner characteristics** | | | | | | | | | | | | |
| **Site** | Caltech | KKI | Leuven | MaxMun | NYU | Olin | Pitt | SDSU | Trinity | UCLA | USM | Yale |
| **Manufacturer, type** | Siemens,  MAGNETOM  TIM Trio | Philips,  Achieva | Philips,  NA | Siemens,  MAGNETOM  Verio | Siemens,  MAGNETOM  Allegra | Siemens,  MAGNETOM  Allegra | Siemens,  MAGNETOM  Allegra | GE,  MR750 | Philips,  Achieva | Siemens,  MAGNETOM  TIM Trio | Siemens,  MAGNETOM  TIM Trio | Siemens,  MAGNETOM  TIM Trio |
| **Field strength** | 3T | 3T | 3T | 3T | 3T | 3T | 3T | 3T | 3T | 3T | 3T | 3T |
| **Headcoil** | NA | NA | NA | NA | NA | NA | NA | 8-ch | 8-ch | NA | NA | NA |
| **T1-weighted image acquisition** | | | | | | | | | | | | |
| **TR/TE/TI (ms)/flip angle (°)** | 1590/  2.73/800/10 | min/  min/1000/8 | min/  4.6/900/8 | 1800/  3.06/900/9 | 2530/  3.25/1100/7 | 2500/  2.74/900/8 | 2100/  3.93/1000/7 | NA/  min  /600/8 | 8.5  3.9/3000/8 | 2300/  2.84/853/9 | 2300/  2.91/900/9 | 1230/  1.73/624/9 |
| **No. of slices** | 176 | 200 | 182 | 160 | 128 | 176 | 176 | 176 | 160 | 160 | 160 | 176 |
| **Voxel size (mm^2^)** | 1.0 x 1.0 | 1.0 x 1.0 | 1.0 x 1.0 | 1.0 x 1.0 | 1.3 x 1.0 | 1.0 x 1.0 | 1.1 x 1.1 | 1.0 x 1.0 | 1.0 x 1.0 | 1.0 x 1.0 | 1.0 x 1.0 | 1.0 x 1.0 |
| **Slice thickness (mm)** | 1.0 | 1.0 | 1.2 | 1.0 | 1.3 | 1.0 | 1.1 | 1.0 | 1.0 | 1.2 | 1.2 | 1.0 |
| **EPI BOLD image acquisition** | | | | | | | | | | | | |
| **Instruction** | Eyes closed | Fixation | Fixation | Fixation or eyes closed | Fixation | Fixation | Eyes closed | Fixation | Eyes closed | Fixation | Eyes open | Eyes open |
| **TR/TE (ms)/flip angle (°)** | 2000/  30/75 | 2500/  30/75 | 1667/  33/90 | 3000/  30/80 | 2000/  15/90 | 1500/  27/60 | 1500/  25/70 | 2000/  30/90 | 2000/  28/90 | 3000/  28/90 | 2000/  28/90 | 2000/  25/60 |
| **No. of volumes** | 150 | 156 | 250 | 120/200 | 180 | 210 | 200 | 180 | 150 | 120 | 240 | 200 |
| **No. of slices** | 34 | 47 | 32 | 40 | 33 | 29 | 29 | 41 | 38 | 34 | 40 | 34 |
| **Voxel size (mm^2^)** | 3.5 × 3.5 | 3.0 × 3.0 | 3.6 × 3.6 | 3.0 × 3.0 | 3.0 × 3.0 | 3.4 × 3.4 | 3.1 × 3.1 | 3.4 × 3.4 | 3.0 × 3.0 | 3.0 × 3.0 | 3.4 × 3.4 | 3.4 × 3.4 |
| **Slice thickness (mm)** | 3.5 | 3.0 | 4.0 | 3.0 | 4.0 | 4.0 | 4.0 | 3.4 | 3.5 | 4.0 | 3.0 | 4.0 |

| **Supplementary Table S14.** Detailed description of MRI acquisition parameters of the respective sites included in ABIDE II. | | | | | | | | | | | | | | |
| --- | --- | --- | --- | --- | --- | --- | --- | --- | --- | --- | --- | --- | --- | --- |
| **Scanner characteristics** | | | | | | | | | | | | | | |
| **Site** | ETH | GU | IU | KKI | KUL | NYU1 | NYU2 | OHSU | ONRC | SDSU | TCD | UCLA | UCD | USM |
| **Manufacturer, type** | Philips,  Achieva | Siemens,  MAGNETOM  TIM Trio | Siemens,  MAGNETOM  TIM Trio | Philips,  Achieva | Philips,  NA | Siemens,  MAGNETOM  Allegra | Siemens,  MAGNETOM  Allegra | Siemens,  MAGNETOM  TIM Trio | Siemens,  MAGNET  OM Skyra | GE,  MR750 | Philips,  Achieva | Siemens,  MAGNETOM  TIM Trio | Siemens,  MAGNETOM  TIM Trio | Siemens,  MAGNETOM  TIM Trio |
| **Field strength** | 3T | 3T | 3T | 3T | 3T | 3T | 3T | 3T | 3T | 3T | 3T | 3T | 3T | 3T |
| **Headcoil** | 32-ch | 12-ch | 32-ch | 8/32-ch | 32-ch | NA | NA | 12-ch | NA | 8-ch | 8-ch | 12-ch | 8-ch | 12-ch |
| **T1-weighted image acquisition** | | | | | | | | | | | | | | |
| **TR/TE/TI (ms)/flip angle (°)** | 8.4/  min/1150  /8 | 2530/  3.5/1100/  7 | 2400/  2.3/1000/  8 | min/  min/1000  /8 | min  4.6/900  /8 | 2530/  3.25/1100  /7 | 2530/  3.25/1100  /7 | 2300/  3.58/900/  10 | 2200/  2.88/794/  13 | NA/  min/600  /8 | 8.4/  3.9/1150  /8 | 2300/  2.86/853/  9 | 2000/  3.16/1050  /8 | 900/  2.91/900/  9 |
| **No. of slices** | 180 | 176 | 256 | 200 | 182 | 128 | 128 | 160 | 208 | 176 | 190 | 160 | 192 | 160 |
| **Voxel size (mm^2^)** | 0.9 x 0.9 | 1.0 x 1.0 | 0.7 x 0.7 | 1.0 x 1.0 | 1.2 x 1.0 | 1.0 x 1.2 | 1.3 x 1.0 | 1.0 x 1.0 | 0.8 x 0.8 | 1.0 x 1.0 | 0.9 x 0.9 | 1.0 x 1.0 | 1.0 x 1.0 | 1.0 x 1.0 |
| **Slice thickness (mm)** | 0.9 | 1.0 | 0.7 | 1.0 | 1.2 | 1.3 | 1.3 | 1.1 | 0.8 | 1.0 | 0.9 | 1.2 | 1.0 | 1.2 |
| **EPI BOLD image acquisition** | | | | | | | | | | | | | | |
| **Instruction** | Fixation | Eyes open | Eyes open | Fixation | Fixation | Fixation | Fixation | Fixation | Fixation | Fixation | Fixation | Fixation | Eyes open | Eyes open |
| **TR/TE (ms)/flip angle (°)** | 2000/  25/90 | 2000/  30/90 | 813/  28/60 | 2500/  30/75 | 2500/  30/90 | 2000/  15/90 | 2000/  30/82 | 2000/  30/90 | 475/  30/60 | 2000/  30/90 | 2000/  27/90 | 3000/  28/90 | 2000/  24/90 | 2000/  28/90 |
| **No. of volumes** | 210 | 154 | 433 | 156 | 162 | 180 | 180 | 120 | 947 | 180 | 210 | 120 | 460 | 240 |
| **No. of slices** | 40 | 43 | 42 | 47 | 45 | 33 | 34 | 36 | 48 | 41 | 37 | 34 | 36 | 40 |
| **Voxel size (mm^2^)** | 3.0 × 3.1 | 3.0 × 3.0 | 3.4 × 3.4 | 3.0 × 3.0 | 2.5 × 2.5 | 3.0 × 3.0 | 3.0 × 3.0 | 3.8 × 3.8 | 3.0 × 3.0 | 3.4 × 3.4 | 3.0 × 3.0 | 3.0 × 3.0 | 3.5 × 3.5 | 3.4 × 3.4 |
| **Slice thickness (mm)** | 3.0 | 2.5 | 3.4 | 3.0 | 2.7 | 4.0 | 3.0 | 3.8 | 3.0 | 3.4 | 3.2 | 4.0 | 4.0 | 3.0 |
